# Elucidating the clinical and molecular spectrum of *SMARCC2*-associated NDD in a cohort of 65 affected individuals

**DOI:** 10.1101/2023.03.30.23287962

**Authors:** Elisabeth Bosch, Bernt Popp, Esther Güse, Cindy Skinner, Pleuntje J. van der Sluijs, Isabelle Maystadt, Anna Maria Pinto, Alessandra Renieri, Lucia Pia Bruno, Stefania Granata, Carlo Marcelis, Özlem Baysal, Dewi Hartwich, Laura Holthöfer, Bertrand Isidor, Benjamin Cogne, Dagmar Wieczorek, Valeria Capra, Marcello Scala, Patrizia De Marco, Marzia Ognibene, Rami Abou Jamra, Konrad Platzer, Lauren B. Carter, Outi Kuismin, Arie van Haeringen, Reza Maroofian, Irene Valenzuela, Ivon Cuscò, Julian A. Martinez-Agosto, Ahna M. Rabani, Heather C. Mefford, Elaine M. Pereira, Charlotte Close, Kwame Anyane-Yeboa, Mallory Wagner, Mark C. Hannibal, Pia Zacher, Isabelle Thiffault, Gea Beunders, Muhammad Umair, Priya T. Bhola, Erin McGinnis, John Millichap, Jiddeke M van de Kamp, Eloise J. Prijoles, Amy Dobson, Amelle Shillington, Brett H. Graham, Evan-Jacob Garcia, Maureen Kelly Galindo, Fabienne G. Ropers, Esther AR Nibbeling, Gail Hubbard, Catherine Karimov, Guido Goj, Renee Bend, Julie Rath, Michelle M Morrow, Francisca Millan, Vincenzo Salpietro, Annalaura Torella, Vincenzo Nigro, Mitja Kurki, Roger E Stevenson, Gijs W.E. Santen, Markus Zweier, Philippe M. Campeau, Mariasavina Severino, André Reis, Andrea Accogli, Georgia Vasileiou

## Abstract

**PURPOSE:** Coffin-Siris and Nicolaides-Baraitser syndromes, are recognisable neurodevelopmental disorders caused by germline variants in BAF complex subunits. The *SMARCC2* BAFopathy was recently reported. Herein, we present clinical and molecular data on a large cohort.

**METHODS:** Clinical symptoms for 41 novel and 24 previously published cases were analyzed using the Human Phenotype Ontology. For genotype-phenotype correlation, molecular data were standardized and grouped into non-truncating and likely gene-disrupting variants (LGD). Missense variant protein expression and BAF subunit interactions were examined using 3D protein modeling, co-immunoprecipitation, and proximity-ligation assays.

**RESULTS:** Neurodevelopmental delay with intellectual disability, muscular hypotonia and behavioral disorders were the major manifestations. Clinical hallmarks of BAFopathies were rare. Clinical presentation differed significantly, with LGD variants being predominantly inherited and associated with mildly reduced or normal cognitive development, while non-truncating variants were mostly *de novo* and presented with severe developmental delay. These distinct manifestations and non-truncating variant clustering in functional domains suggest different pathomechanisms. *In vitro* testing showed decreased protein expression for N-terminal missense variants similar to LGD.

**CONCLUSION:** This study improved *SMARCC2* variant classification and identified discernible *SMARCC2*-associated phenotypes for LGD and non-truncating variants, which were distinct from other BAFopathies. The pathomechanism of most non-truncating variants has yet to be investigated.

## INTRODUCTION

The BAF (BRG1/BRM-associated factor) complex is an ATP-dependent chromatin remodeling complex that repositions nucleosomes and increases the accessibility of regulatory DNA sequences.^1^ “BAFopathies” encompass a spectrum of neurodevelopmental delay disorders (NDDs) caused by germline pathogenic variants in BAF complex subunit genes including *SMARCA4*, *SMARCA2*, *ARID1A/B*, *SMARCB1/E1*, *DPF2*, and *ARID2*. The most well-defined BAFopathies with overlapping clinical presentations are Coffin-Siris (CSS; MIM 135900) and Nicolaides-Baraitser (NCBRS; MIM 601358) syndromes. ^2–7^

Recent studies reporting affected individuals with mainly *de novo* missense/in-frame and a few likely gene-disrupting (LGD) pathogenic variants in another BAF-subunit, *SMARCC2* (*BAF170*, MIM *601734), expanded the spectrum of BAF-related NDDs. ^8–13^ Machol *et al.* described a cohort of 15 cases with variable clinical manifestations resembling CSS and NCBRS. ^8^ The Online Mendelian Inheritance in Man (OMIM) database now classifies the *SMARCC2*-associated phenotype as Coffin-Siris syndrome 8 (MIM #601734). The described phenotype included neurodevelopmental delay (DD), mild to severe intellectual disability (ID), profound speech delay, behavioral abnormalities, muscular hypotonia, feeding disorders in infancy, and dysmorphic facial features.

SMARCC2 contains four well described and highly conserved functional domains, namely the SWIRM domain, the SANT domain, and two domains in a coiled-coil region, termed dimerization (DR) and core assembly region (CAR) (see Figure 1A). Constitutional abolition of Smarcc2 during postnatal and adult hippocampal neurogenesis in mice increased astrogenesis, resulting in an abnormal spatial distribution of radial glial-like cells, ultimately linked to behavioral and learning impairments. ^14^ Additionally, intact Smarcc2 expression determines cerebral cortex volume, thickness, forebrain and cortex development. ^14, 15^

**Figure 1.**
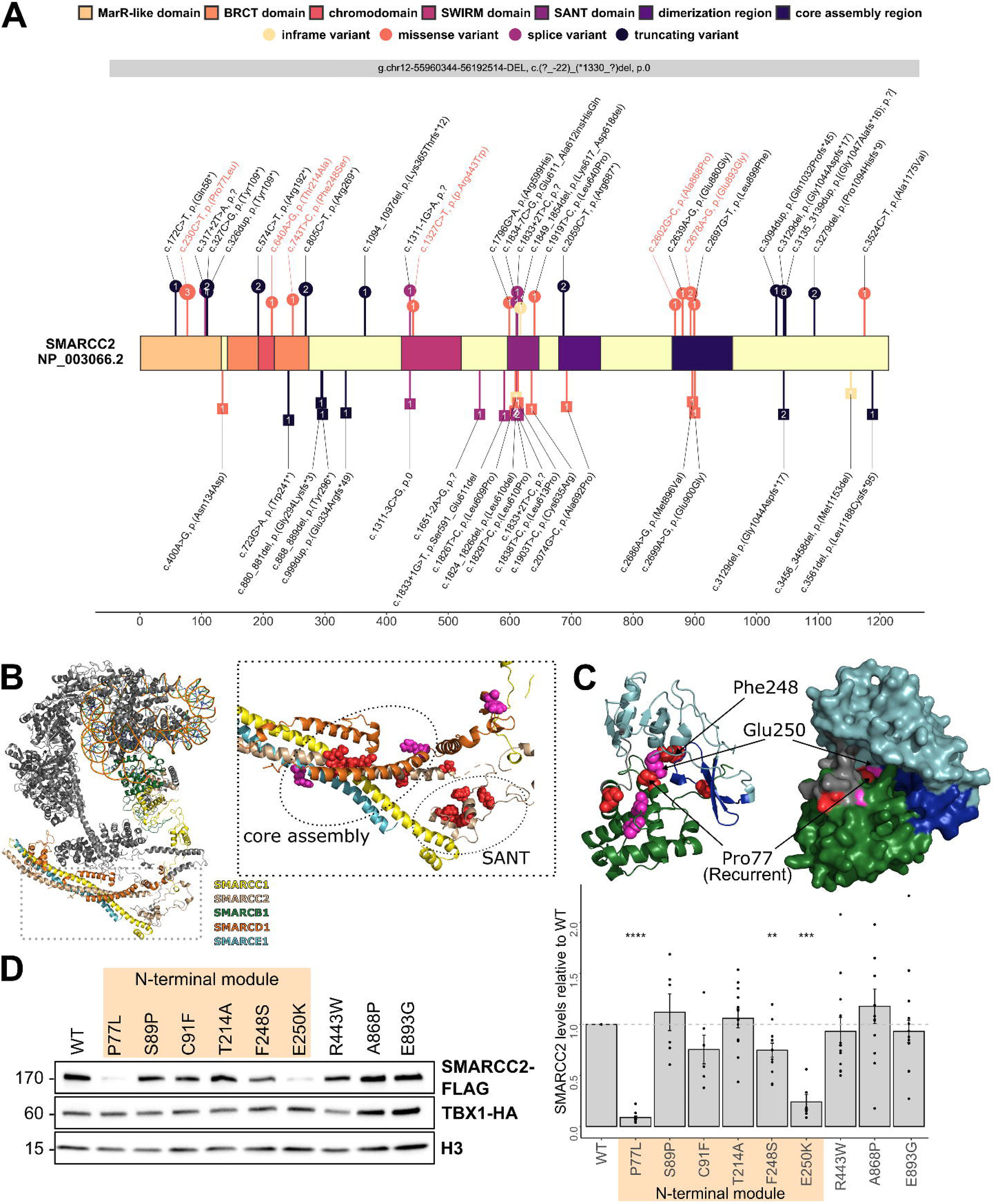
SMARCC2 linear domain structure, distribution of pathogenic variants in the BAF complex and N-terminal variants causing protein loss. (A) Linear protein model of SMARCC2 and its domains: an N-terminal module containing a MarR-like helix-turn-helix domain (eggshell) with DNA-binding ability ^26^, as well as a BRCT domain (orange) with an inserted non-functional chromodomain (red), which have been proposed to mediate protein-protein interactions. ^26^ The SWIRM domain (magenta) mediates protein-protein interaction, ^35, 36^ the SANT domain (berry) is the chromatin binding domain of the protein which was proposed to recognize unmodified histone tails, ^37, 38^ the dimerization region (purple) and core assembly region (dark blue) are coiled-coil domains involved in the formation of the core BAF complex. The first is necessary for heterodimerization with SMARCC1 and the latter interacts with SMARCD1 and SMARCE1, forming the base of the BAF core module ^24^. Circles above the protein model indicate novel heterozygous variants, while squares below the model indicate previously described variants (beige: inframe; orange: missense; berry: splice; black: truncating). The size of the lollipop represents the number of individuals with the variant, the number inside shows the number of families. The variant’s scaled CADD (Combined Annotation Dependent Depletion) scores ^39^ are the lollipop segment length. Orange labelled variants were used for functional analysis. (B) Left side with an overview of the BAF complex as a cartoon model (based on PDBDEV_0000556). The BAF subunits constituting the base (core) module are colored according to the legend, whereas the other subunits are shown in gray. Right side with a magnification of the base module displaying the fingers submodule ^25^ with the characteristic five-helix bundle formed by domains of SMARCC1, SMARCC2, SMARCD1 and SMARCE1 and the SANT domain of SMARCC2 constituting the “palm” submodule. The missense variants in SMARCC2 are shown in red, whereas the published variants in other base modules are shown in magenta. The core assembly and SANT domains of SMARCC2 are denoted by ellipses and contain an appreciable cluster of missense variants in close proximity, emphasizing their functional significance. The close proximity of the variants in the core assembly region to the alpha helices from other subdomains and variants in these (SMARCE1: p.(Arg251Gln); SMARCD1: p.(Arg446Gly)) suggests that the interaction between these and thus the formation of the base scaffold may be compromised. (C) The homology model of the N-terminus of SMARCC2 with the MarR-like (green), BRCT (cyan), and chromodomain (blue) domains is depicted as a cartoon on the left. The SMARCC2 missense variants identified in this cohort are depicted in red, while synthetic variants previously identified in tumors ^26^ are depicted in magenta. The right side depicts a surface rendering of the homology model, which reveals that the p.(Pro77Leu) hotspot variant is in close proximity to p.(Phe248Ser) and the cancer-related mutation p.Glu250Lys. The other variants are distributed throughout the globular N-terminus. Missense3D ^32^ predicts that both p.(Pro77Leu) and p.Glu250Lys are structurally damaging. (D) SMARCC2 protein expression. Left panel: representative western blot image. TBX1-HA was used as transfection control, Histone H3 as loading control. Right panel: quantification of SMARCC2 protein levels. Wildtype and mutant SMARCC2-FLAG was normalized to TBX1-HA and the value of wildtype SMARCC2-FLAG was set to 1 (marked by dashed line). Data stems from at least 6 independent experiments. Sample means are depicted as bars with SEM, individual values as dots. P-values were calculated using a one sample t-test (hypothetical mean = 1, significance threshold < 0.05). **** p < 0.0001, *** p < 0.001. ** p < 0.01. Note significant protein expression loss for N-terminal variants c.230C>T p.(Pro77Leu), c.743T>C p.(Phe248Ser), and c.748G>A p.(Glu250Lys).

To date, reports of *SMARCC2* variants have been mostly part of individual case reports or large NDD studies with limited clinical information. ^9–13, 16^ In an attempt to better characterize the clinical and molecular spectrum of *SMARCC2-*associated

NDD, a large cohort of 65 cases was collected, including 41 novel individuals with *de novo* or inherited variants, whose clinical and molecular findings were systematically described, and 24 previously published individuals, whose data were thoroughly curated. Additionally, Human Phenotype Ontology (HPO) and automated facial recognition were used to investigate genotype-phenotype correlations between non-truncating and LGD variants, and structural modeling as well as functional assays to investigate missense variants.

## MATERIALS AND METHODS

### Cohort and ethical considerations

A cohort of 65 individuals with *SMARCC2* variants, including 24 previously reported and 41 novel, was collected (File S1 and S2). Two individuals (Ind-18; c.172C>T p.(Gln58*), Ind-25; c.1094_1097del p.(Lys365Thrfs*12)) previously described in other publications, albeit with incomplete clinical or molecular characterization, were included in the novel series. ^9, 10^ Novel *SMARCC2* individuals were recruited using GeneMatcher ^17^ and an international collaborative network. This study follows the Declaration of Helsinki. Genetic testing was done in routine diagnostic settings (n = 30) or in research settings (n = 11) after ethical review board approval. Legal guardians gave written informed consent for genetic and clinical data, including photos and brain images, to be published. See File S2 sheet “clinical_table” for setting.

### Genetic analysis

The majority of *SMARCC2* variants was found by exome sequencing (singleton n=14, duo n=4 and trios n=20) in the collaborating centers using different analysis platforms based on BWA/GATK pipelines. ^18, 19^ Chromosomal microarray revealed a complete *SMARCC2* gene deletion in Ind-12. RT-PCR for Ind-29 was performed using standard methods. Real-time PCR was used to measure *SMARCC2* expression levels of Ind-19 (Fam-18). See File S2 sheet “clinical_table” for genetic analyses and File S1 for method details.

### Variant annotation and scoring

Variants were standardized to the *SMARCC2* reference transcript NM_003075.5 (GRCh37/hg19) using Mutalyzer 3 ^20^ and annotated using the Ensembl Variant Effect Predictor (VEP) ^21^. All *SMARCC2* variants were subsequently reclassified based on the recommendations of the American College of Medical Genetics and Genomics (ACMG) ^22^ and subsequent updates. An *a priori* and *a posteriori* classification system was used based on either prior evidence or our findings, such as new mutational hotspots, clustering in functional domains, recurrence, and functional results supporting pathogenicity. Compare File S2 sheets “clinical_table” and “ACMG criteria”.

### Clinical Information

Clinical manifestations were systematically described and standardized according to the HPO terminology (File S2 sheet “clinical_table” and File S1 “Clinical reports”). ^23^ Information on clinical abnormalities and facial dysmorphic features, which were available for 58 *SMARCC2* subjects, including all novel ones, were summarized and compared (Table 1 and 2, Table S1). When available, cranial magnetic resonance imaging (MRI) data were evaluated by an experienced pediatric neuroradiologist. The facial overlays shown in Figure 2 were generated by applying the Face2Gene research application (FDNA Inc., Boston, MA, USA) to a total of 23 novel and previously published *SMARCC2* individuals (Figure 2 and File S1 “Clinical information”).

**Figure 2.**
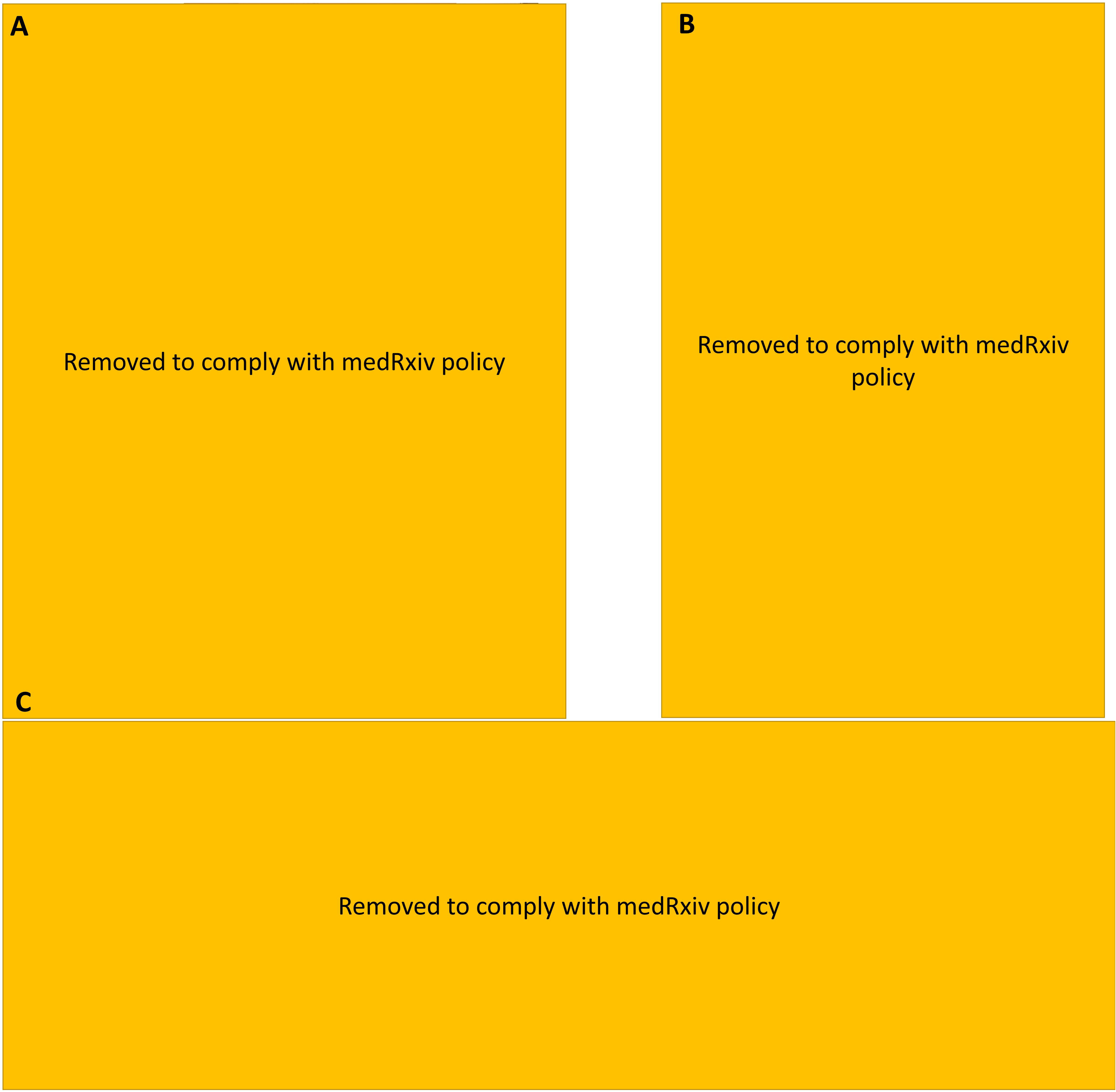
Facial appearance and representative cMRIs of *SMARCC2* individuals. Facial features, facial overlay and images from hands and feet of individuals with LGD **(A)** and non-truncating **(B)** *SMARCC2* variants. Ind-7 and Ind-8 carrying the missense variant c.230C>T p.(Pro77Leu), which leads to SMARCC2 protein loss, were grouped together with the carriers of LGD variants. Note the pronounced coarseness of facial characteristics in individuals with non-truncating variants. (**C**) Neuroimaging characteristics of *SMARCC2* individuals and a control subject. Ind-02, Ind-09 and Ind-11 exhibit normal corpus callosum appearance. Corpus callosum hypoplasia (empty arrows) or dysplasia with thinning of the corpus callosum splenium (thick arrows), may or may not be accompanied by anterior commissure hypoplasia/agenesis (arrowheads) and small inferior cerebellar vermis (thin arrows).

**Table 1.**
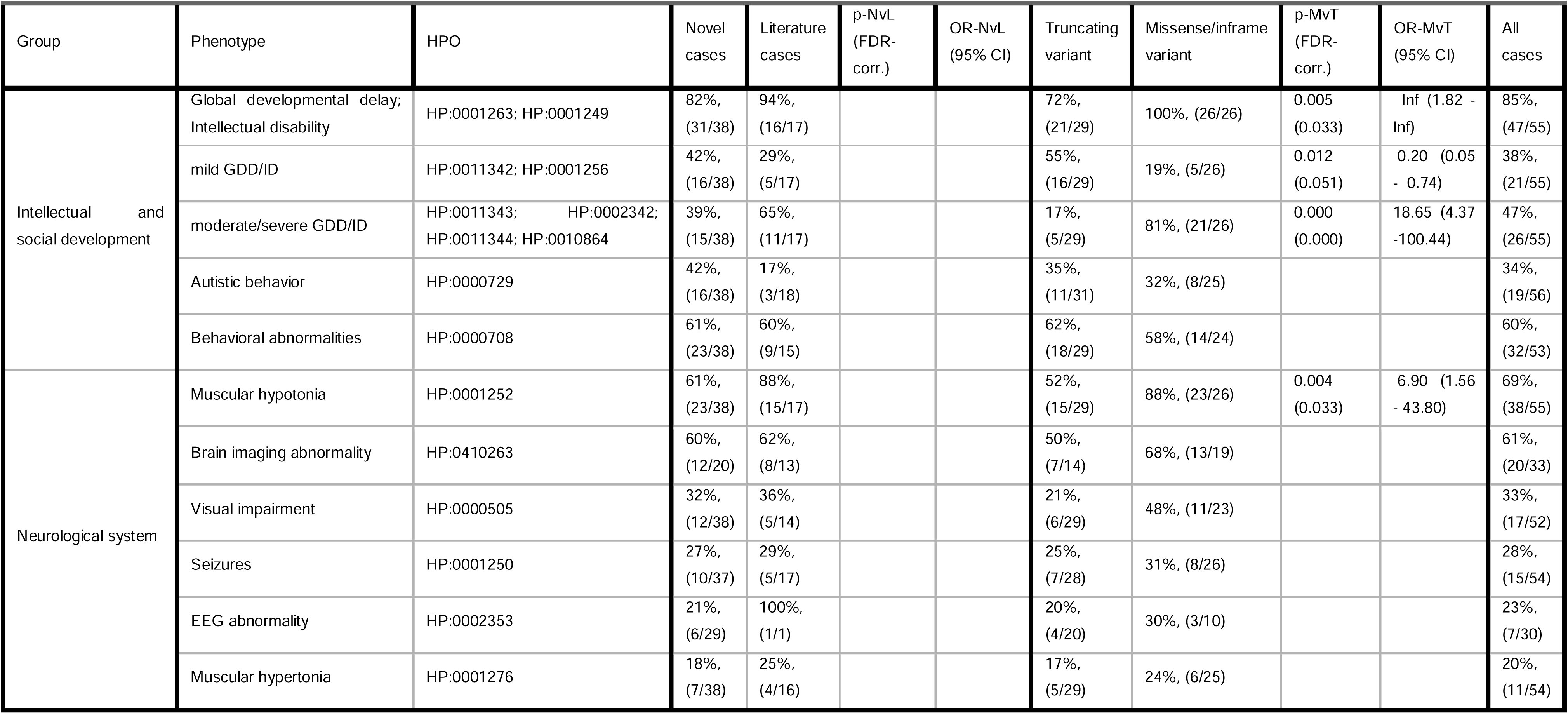
Neurodevelopmental and neurological phenotypes. p-NvL: p-value novel versus literature cohorts; OR-NvL: odds ratio novel versus literature cohorts; p-MvT: p-value missense versus truncating variant cohorts; OR-MvT: odds ratio missense versus truncating variant cohorts; FDR-corr.: corrected false discovery rate; CI: confidence interval

| Group | Phenotype | HPO | Novel cases | Literature cases | p-NvL (FDR-corr.) | OR-NvL (95% CI) | Truncating variant | Missense/inframe variant | p-MvT (FDR-corr.) | OR-MvT (95% CI) | All cases |
| --- | --- | --- | --- | --- | --- | --- | --- | --- | --- | --- | --- |
| Intellectual and social development | Global developmental delay; Intellectual disability | HP:0001263; HP:0001249 | 82%, (31/38) | 94%, (16/17) |  |  | 72%, (21/29) | 100%, (26/26) | 0.005 (0.033) | Inf (1.82 - Inf) | 85%, (47/55) |
|  | mild GDD/ID | HP:0011342; HP:0001256 | 42%, (16/38) | 29%, (5/17) |  |  | 55%, (16/29) | 19%, (5/26) | 0.012 (0.051) | 0.20 (0.05 - 0.74) | 38%, (21/55) |
|  | moderate/severe GDD/ID | HP:0011343; HP:0002342; HP:0011344; HP:0010864 | 39%, (15/38) | 65%, (11/17) |  |  | 17%, (5/29) | 81%, (21/26) | 0.000 (0.000) | 18.65 (4.37 - 100.44) | 47%, (26/55) |
|  | Autistic behavior | HP:0000729 | 42%, (16/38) | 17%, (3/18) |  |  | 35%, (11/31) | 32%, (8/25) |  |  | 34%, (19/56) |
|  | Behavioral abnormalities | HP:0000708 | 61%, (23/38) | 60%, (9/15) |  |  | 62%, (18/29) | 58%, (14/24) |  |  | 60%, (32/53) |
| Neurological system | Muscular hypotonia | HP:0001252 | 61%, (23/38) | 88%, (15/17) |  |  | 52%, (15/29) | 88%, (23/26) | 0.004 (0.033) | 6.90 (1.56 - 43.80) | 69%, (38/55) |
|  | Brain imaging abnormality | HP:0410263 | 60%, (12/20) | 62%, (8/13) |  |  | 50%, (7/14) | 68%, (13/19) |  |  | 61%, (20/33) |
|  | Visual impairment | HP:0000505 | 32%, (12/38) | 36%, (5/14) |  |  | 21%, (6/29) | 48%, (11/23) |  |  | 33%, (17/52) |
|  | Seizures | HP:0001250 | 27%, (10/37) | 29%, (5/17) |  |  | 25%, (7/28) | 31%, (8/26) |  |  | 28%, (15/54) |
|  | EEG abnormality | HP:0002353 | 21%, (6/29) | 100%, (1/1) |  |  | 20%, (4/20) | 30%, (3/10) |  |  | 23%, (7/30) |
|  | Muscular hypertonia | HP:0001276 | 18%, (7/38) | 25%, (4/16) |  |  | 17%, (5/29) | 24%, (6/25) |  |  | 20%, (11/54) |

Fisher’s exact test was used to calculate p-values for novel cases compared to reviewed and published cases. Multiple testing was adjusted for using false discovery rate (FDR with threshold < 0.05). Comparing missense/in-frame and LGD causative carrier groups followed the same method. All computations were done in R 4.1.3 using RStudio (Table 1 and 2, Table S1).

### Protein model analysis of missense variants

We used the published structures PDBDEV_00000056 ^24^ (PDB-Dev) and 6LTH ^25^ (protein data bank; PDB) to map disease associated variants of BAF complex subunits. We utilized the published crystal structure of the SMARCC1 N-terminus (6YXO ^26^) to generate a homology model (File S4) of the paralogous SMARCC2 region with PHYRE2 ^27^ (One-to-One threading option), which we used to investigate disease associated variants in this region. Structures were visualized with the Pymol software (OpenSource Version 2.5.0; Schrodinger, LLC). Missense variants in other BAF base complex subunits (File S2 sheet “missense_other_BAF”) were reviewed from published literature reports (Table S2), standardized to fit the model transcript and used to analyze spatial proximity in the BAF complex model.

### Functional analyses of missense variants

FLAG-tagged SMARCC2 was obtained from Addgene (plasmid #19142) ^28^ and variants were introduced using the In-Fusion HD Cloning Kit (Clontech). Seven mutants harboring missense variants in different protein domains were generated: p.(Pro77Leu), p.(Thr214Ala) and p.(Phe248Ser) in the N-terminal module, p.(Arg443Trp) in the SWIRM domain, p.(Leu640Pro) in the SANT domain and p.(Ala868Pro) and p.(Glu893Gly) in the CAR. Plasmids were transfected into HEK293T cells using JetPrime (Polyplus Life Science). Immunofluorescence staining was performed as previously described. ^29^ Proximity ligation assay (PLA) was performed using Duolink In Situ Reagents (Sigma). Co-immunoprecipitation (CoIP) was carried out as previously described. ^30^ Protein stability was assessed by transiently co-expressing FLAG-tagged SMARCC2 variants together with a different-sized control protein (HA-tagged TBX1), followed by quantitative western blot analysis and normalization of SMARCC2-FLAG to TBX1-HA. File S1 “Supplemental methods” contain experimental details, oligonucleotide sequences (Table S4), and antibodies (Table S5).

## RESULTS

### Description of *SMARCC2* variants

Of the 45 *SMARCC2* variants described in this study, 25 are novel. Two alterations have been reported in both this and previous studies. ^8, 12^ Three novel missense substitutions (c.2697G>T p.(Leu899Phe), c.640A>G p.(Thr214Ala), and c.1327C>T p.(Arg443Trp)) and one previously described splice variant (c.1651-2A>G p.?) ^13^ could not be further classified due to lack of conclusive evidence (no strong segregation information, recurrence, or strong effects in functional studies). Thus, carriers Ind-03, Ind-06, Ind-40, and Gofin_Subject 5 were excluded from further clinical analysis. The linear model (Figure 1A) shows the variants, and File S2 sheet “clinical_table” and File S1 “clinical reports” describe these individuals clinically.

The present study describes 27 probably non-truncating variants, including 19 missense (11 novel), three in-frame (one novel), and five splice (two novel). All non-truncating variants arose *de novo*. Segregation could not be determined for the splice changes c.1651-2A>G and c.1833+1G>T. The splice variants c.1311-1G>A, c.1651-2A>G and c.1833+2T>C were computationally predicted (http://autopvs1.genetics.bgi.com^31^) to result in an in-frame deletion. RNA samples were unavailable for further analysis. The splice donor variant 1833+1G>T causes an in-frame deletion of exon 19, creating an aberrant product that escapes NMD. ^8^ Messenger RNA sequencing for the novel intronic variant c.1834-7C>G revealed an aberrant transcript with retention of the last 6 bp of intron 19 (r.1833+1_1834-1_ins1834-6) and an in-frame insertion of two amino acids (p.Glu611_Ala612insHisGln) that was less stable (Figure S4A-C).

Seven previously reported missense/in-frame causative variants clustered in the highly conserved SANT domain. Variant c.1833+2T>C has been reported in two unrelated individuals in a previous study ^8^ and once in the present study. Each of the amino acid substitutions c.1829T>C p.(Leu610Pro) and c.1826T>C p.(Leu609Pro) was detected in two different families in previous studies. The clustering of pathogenic variants in this functional domain was confirmed by the discovery of four novel non-truncating variants, two missense and two in-frame. Consequently, *de novo* missense alterations in the SANT domain could be classified *a priori* as likely pathogenic (PS2_Supporting, PM2_Supporting, PP2_Supporting, PM1_Moderate) upon meeting the cutoffs for computational evidence (PP3).

Machol *et al.* ^8^ reported two non-truncating variants in the CAR domain, here classified as a clustering hotspot due to four new amino acid substitutions, one of which was found in two unrelated families (c.2678A>G p.(Glu893Gly)). *De novo* non-truncating variants in this domain were classified as likely pathogenic according to the PM1_Moderate criterion if computational predictions (CADD PHRED v1.6 score ≥28.1) indicated they were pathogenic. Variants in the DR and SWIRM domains are rare and lack molecular/functional evidence, thus classified as variants of unknown significance (VUS). N-terminal missense variants included one previously reported ^8^ and three novel ones. The substitution c.230C>T p.(Pro77Leu) in three unrelated families was *post hoc* interpreted as likely pathogenic after considering variant recurrence and functional assays. The remaining N-terminal and variants outside of SMARCC2 domains remained VUS due to lack of evidence.

Overall, the 18 identified LGD variants were dispersed across *SMARCC2.* Seven of these variants have been previously reported (two nonsense, four frameshifting, and one splice). The remaining 11 variants were novel consisting of six nonsense, three frameshifting, and two splice. Most LGD alterations were inherited from unaffected parents. Four LGD variants were *de novo*, and seven individuals had unknown inheritance patterns. All LGD variants could be classified as (likely) pathogenic (PVS1_VeryStrong, PM2_Supporting) assuming loss-of-function (LOF) as disease mechanism. Ind-12 had a microdeletion that included *SMARCC2* and 14 other genes, including *RPS26*, which is linked to dominantly inherited Diamond-Blackfan anemia 10 (MIM #613309). The splice variants c.317+2T>A p.? and c.3135_3139dup p.[(Gly1047Alafs*16); p.?] were computationally predicted to cause out-of-frame effects. Real-time PCR previously demonstrated that the variant c.1311-3C>G reduces *SMARCC2* expression. ^8^ Variants c.327C>G p.(Tyr109*), c.574C>T p.(Arg192*), c.805C>T p.(Arg269*), c.2059C>T p.(Arg687*) were each found in two unrelated individuals and variant c.3279del p.(Pro1094Hisfs*9) in two siblings in the novel cohort. Variant c.3129del p.(Gly1044Aspfs*17) was prior reported in a twin pair ^12^ as well as identified in five siblings and one unrelated individual in the present cohort. Also, variant c.3129del p.(Gly1044Aspfs*17) was the only LGD variant of the combined *SMARCC2* cohort to be listed in gnomAD (seven heterozygous carriers). We used the well characterized GeneDx cohort to examine the prevalence of this variant in individuals with DD/ID (10/97.993) and healthy controls (13/400.200). The odds-ratio (OR) was calculated using Fisher’s exact test and showed a moderate to strong association (OR: ∼ 3.14, p-value < 0.008, 95% CI: 1.23 - 7.76). Real-time PCR on peripheral blood from one Fam-18 carrier (Ind-19) confirmed NMD with reduced *SMARCC2* expression level at nearly 68% (Figure S4D).

Three individuals (8.5% of this cohort) carried a second (likely) pathogenic variant in an NDD-related gene (Ind-25, Ind-33, Ind-34) (see also File S1 “Supplemental Results” and File S2 sheet “clinical_table” for these and additional VUS).

### Linear and 3D protein model

Annotation of variants on the linear gene model revealed that truncating variants were dispersed throughout *SMARCC2*, whereas non-truncating variants clustered within the SANT and core assembly domains (Figure 1A). Furthermore, a correlation between these clusters and high computational prediction scores for missense variants in the annotated domains was found (Figure S1). Mapping of the amino acid residues to the three-dimensional protein structure of the BAF complex, demonstrated that missense variants in the core assembly domain are located in the five-helix bundle of the base module, which potentially impedes interaction with other subunits forming the base scaffold (Figure 1B). The homology model revealed that missense variants in the N-terminal region, which is not covered in any of the two published BAF complex structures ^24, 25^, are generally dispersed throughout the globular domain, except variants p.(Pro77Leu) and p.(Phe248Ser) which are found in close proximity to each other, as well as the cancer-related variant p.Glu250Lys (Figure 1C). Exclusively the p.(Pro77Leu) variant in the N-terminus was predicted to cause structural changes using Missense3D ^32^ (File S3 sheet “cohort_variants”).

### Functional analysis of SMARCC2 missense variants

As missense variants in other BAF subunits have been linked to protein misfolding and aggregate formation, we initially investigated cellular SMARCC2 protein localization in a subset of missense alterations, but all of them exhibited normal nuclear localization (Figure S5). We next analyzed whether the missense variants affect the BAF complex combinatorial assembly. Both PLA and quantitative CoIP showed no impairment of SMARCC2 interaction with the subunits ARID1B, SMARCA4, SMARCC1 and SMARCE1 (Figure S6). Co-immunoprecipitation experiments showed a trend towards higher interaction of mutant SMARCC2-FLAG with SMARCC1 as compared to the wildtype protein, however this effect did not reach statistical significance. Finally, we asked whether *SMARCC2* variants affect protein stability. We included three cancer-related missense alterations (p.(Ser89Pro), p.(Cys91Phe), p.(Glu250Lys)) located in close proximity to the herein identified N-terminal variants. These were computationally predicted to cause structural defects of the SMARCC2 protein in a previous study. ^26^ While variants in other protein regions did not adversely affect protein stability, four of the investigated N-terminal variants showed an impact: the recurrent variant p.(Pro77Leu) from this cohort and the cancer variant p.(Glu250Lys) significantly reduced protein levels (> 80%). Likewise, cohort variant p.(Phe248Ser) and cancer variant p.(Cys91Phe) each resulted in a 20% reduction of protein levels, although only the former was statistically significant (Figure 1D).

### Clinical presentation of *SMARCC2* individuals

Global developmental delay and/or intellectual disability (ID) was described in 85% of *SMARCC2* individuals, with 38% being mildly and 47% moderately/severely affected, whereas 15% had no cognitive or speech/motor deficits. Gross motor delay and fine motor deficits in infancy and late childhood were reported in 55% of cases. Muscular hypotonia was found in 69% of the individuals, and behavioral abnormalities in 60%, with autistic behavior being the most common (34%). Anxiety, aggression, attention deficit hyperactivity disorder (ADHD), fixations, tantrums, and obsessive-compulsive behaviors were also common. Generalized, tonic, tonic-clonic, focal, absence seizures, and Lennox-Gastaux syndrome were diagnosed in 28% of the cohort. Visual defects, primarily due to refraction anomalies such as hypermetropia, hyperopia, astigmatism and myopia were present in 33% of the individuals. Strabismus and ptosis were the most common structural eye abnormalities (27%). Neuroimaging studies in 34 subjects (brain MRI in 32 and CT in two) revealed abnormalities in 21 individuals (61%). Neuroimaging features included non-specific white matter signal alterations, intracranial arachnoid cysts and/or small inferior cerebellar vermis (34%), corpus callosum hypoplasia and/or dysplasia (23%), white matter volume loss and/or anterior commissure agenesis/hypoplasia (14.7%) and enlargement of cerebrospinal fluid spaces (14.7%) (Figure 2C, File S2 sheet “clinical_table”, and Figure S3).

Seven individuals had low birth weight and length, while seven had high birth weight and normal length. Two individuals showed oligohydramnios and six intrauterine growth retardation. Reduced body weight and short stature were found in 30% and 24% of the individuals, respectively. Clinodactyly, camptodactyly, brachydactyly, long fingers, and persistent fingertip pads were found in 31% of subjects. Notably, prominent interphalangeal joints were rare (4%) and absent phalanges of the 5th finger were not reported. Only Ind-43 presented with shorter distal phalanx and hypoplastic nail of the left thumb.

Pes planus was the most common foot deformity (29%), while only two individuals had hypoplastic toenails (4%). Ectodermal anomalies such as sparse/thin scalp hair and hypertrichosis were noticed in 16% and 17% of individuals, respectively. The most common skeletal malformation was scoliosis (28%). Aside from genitourinary (23%) and gastrointestinal abnormalities (22%), other congenital disorders, such as heart defects, were relatively uncommon. Feeding difficulties or failure to thrive were reported by half of the cohort (51%). Finally, 21% of *SMARCC2* individuals reported sleep disturbances, and 16% had recurrent infections (Table 2 and Table S1).

**Table 2.**
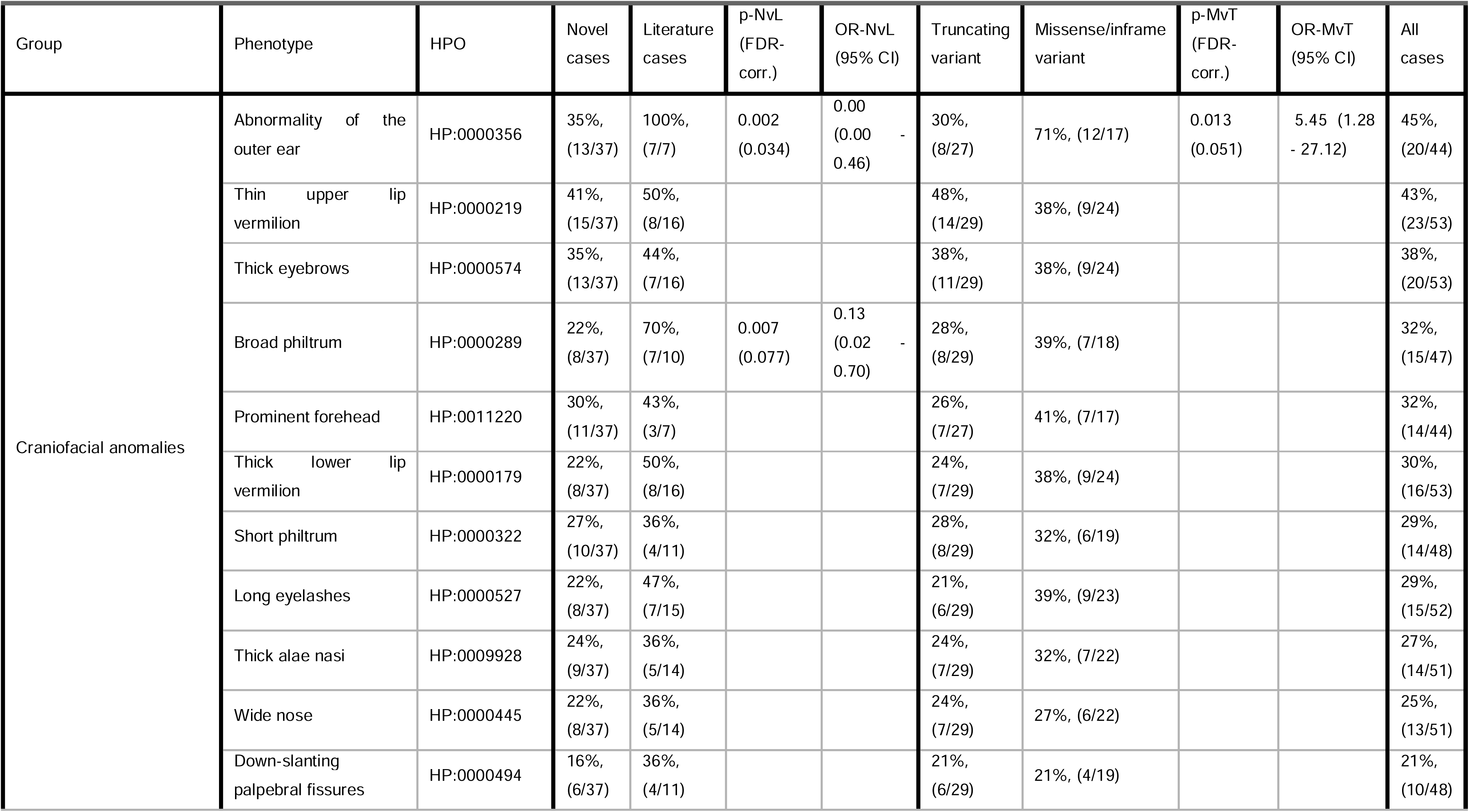
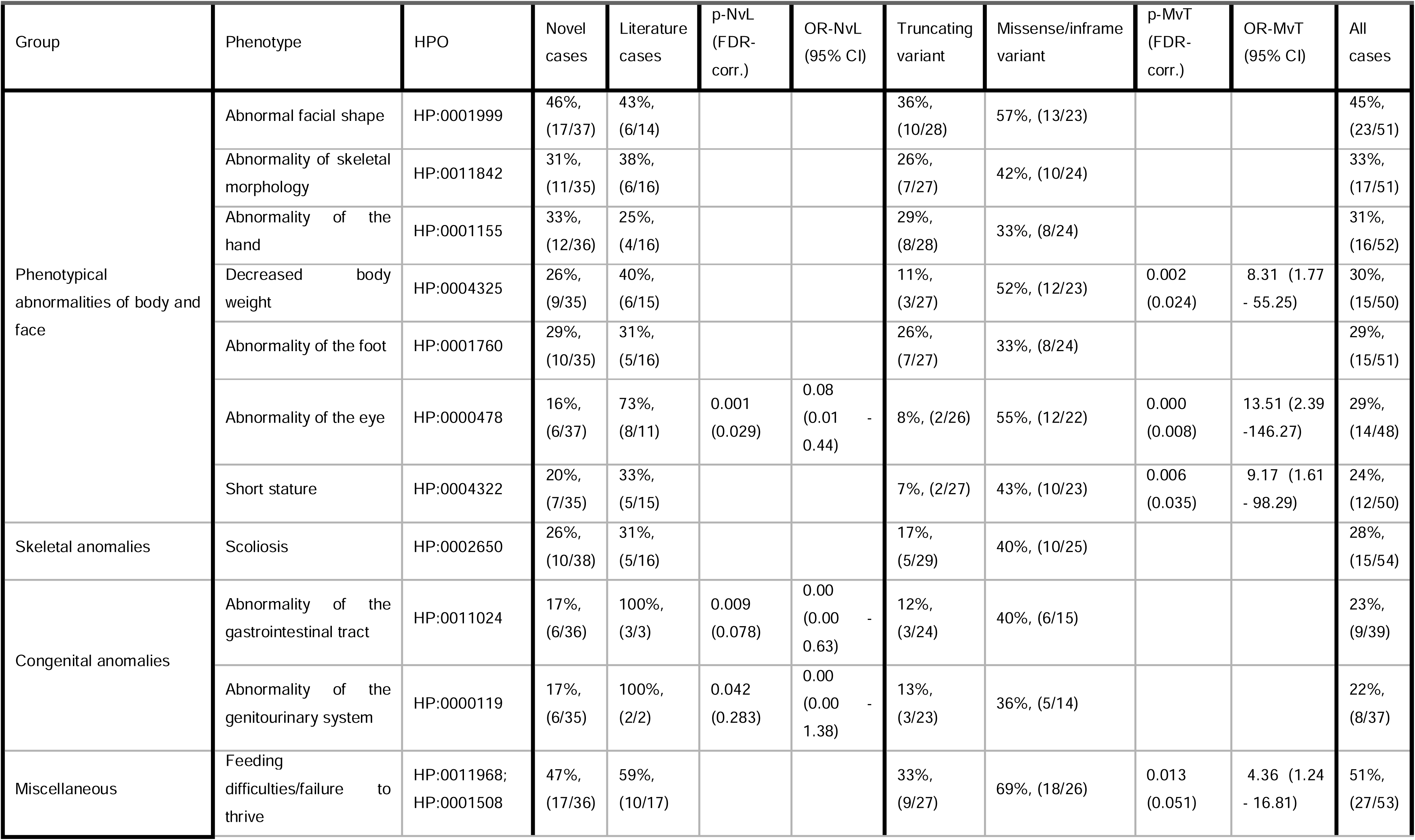
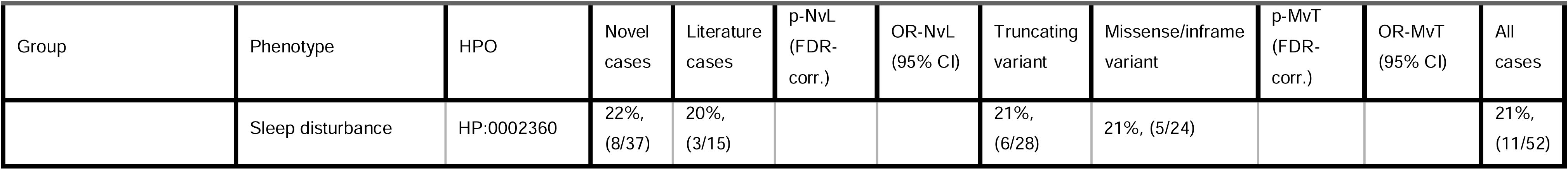
Other phenotype categories. p-NvL: p-value novel versus literature cohorts; OR-NvL: odds ratio novel versus literature cohorts; p-MvT: p-value missense versus truncating variant cohorts; OR-MvT: odds ratio missense versus truncating variant cohorts; FDR-corr.: corrected false discovery rate; CI: confidence interval

Detailed dysmorphic facial features were observed in 45% of *SMARCC2* subjects (Figure 2A and 2B, Table 2, Table S1, and File S2 sheet “clinical_table”). Apart from structural eye and outer ear anomalies, the prevalence of clinical and dysmorphic features between previous studies and this report was consistent (see also Table 1 and 2, File S1 “Supplemental results”).

### Genotype-phenotype correlation in LGD and non-truncating variants

DD/ID was found in 100% of missense/in-frame and 72% of LGD variant carriers. Missense/in-frame variants were associated with moderate/severe DD/ID (81% vs. 17%) with severe speech deficits and normal to moderate motor development. LGD individuals showed predominantly mild DD (55% vs. 19%) with mild/borderline ID or normal cognitive development, mild expressive/receptive language deficits or unaffected speech development, and motor abilities that were normal to mildly restricted in the majority of the cases. Notably, behavioral disorders occurred at equal frequency in both groups. Missense/in-frame variants more frequently caused muscular hypotonia (88% vs 52%). Compared to LGD carriers, missense/in-frame variant carriers had shorter stature (43% vs. 7%) and lower body weight (52% vs. 11%). Structural eye (55% vs. 8%) and outer ear (71% vs. 30%) abnormalities, were more common in individuals with non-truncating alterations (Table 1 and 2, Table S1). Facial recognition revealed a facial gestalt, which was dominated by coarse facies in missense/in-frame carriers (Figure 2A and 2B).

## DISCUSSION

The present study provides novel insight into the *SMARCC2*-associated phenotype, via a comprehensive analysis of a large cohort of individuals with *SMARCC2* variants, both known and novel. Functional assays revealed reduced protein expression as a pathomechanism in a subset of *SMARCC2* N-terminal missense variants. Moreover, the systematic characterization of clinical traits suggests that non-truncating and LGD variants are associated with clinical entities of variable severity.

*SMARCC2* is highly intolerant to non-truncating (Z-score of 3.91) and LGD variants (pLI score of 1 in gnomAD). We found that *de novo* missense/in-frame variants clustered mainly in the SANT and the C-terminal CAR domains, enabling their classification as (likely) pathogenic. SWIRM and DR domain variants are rare. Nevertheless, considering that computational analysis indicated high conservation and constraint of amino acid substitutions for both regions (Figure S1), future studies addressing non-truncating changes in these domains could help identify additional clusters.

All LOF changes were (likely) pathogenic. We focused on the pathogenicity of the c.3129del p.(Gly1044Aspfs*17), as this variant was the only one listed in public databases, and exceeded the allele frequency cutoff (2 in gnomAD). RNA expression analysis in one carrier showed the expected NMD, although it was incomplete. A suspected milder impact of this alteration due to residual expression requires protein analysis, but no further material from this individual or other LGD variant carriers was available to compare expression levels. In the well-studied GeneDx cohort, a moderate to strong effect of this frameshifting variant on DD/ID risk (OR ∼ 3.14, p-Fisher < 0.008) was found. To exclude the possibility that milder effects of this variant confounding the genotype-phenotype analysis, clinical data were re-analyzed after excluding all eight carriers of this alteration in this cohort, and did not detect any deviations as compared to the previous analysis of the non-truncating/LGD group (Table S3). This suggests that the effect of this variant does not differ from that of other LGD counterparts.

A subset of *SMARCC2* variants was dispersed across the N-terminal module, whose function is not well defined to date. A recent study on the SMARCC2 ortholog SMARCC1 found several structural domains in its N-terminus, including a MarR-like helix-turn-helix, chromo-, and BRCT domain, which regulate transcription, histone modifications, and complex activity, respectively. ^33, 34^ Variants in this module were predicted to cause protein folding defects and destabilization. ^26^ SMARCC2’s N-terminal structure shares 66% homology with SMARCC1, suggesting a similar domain composition. 3D protein modeling suggested a significant alteration (contraction of the cavity encompassed by the BRCT and N-terminal domains) only for the recurrent variant p.(Pro77Leu) in this cohort. Functional analysis confirmed the pathogenicity of p.(Pro77Leu) by showing significantly reduced protein stability and almost complete SMARCC2 protein loss, similar to LGD variants. Notably, the synthetic variant p.(Glu250Lys), previously suggested to cause a structural defect, ^26^ resulted in a similar effect, while the variant p.(Phe248Ser) in this cohort, which is structurally close to both, had a weaker effect. These findings emphasize the structural importance of the N-terminal region without, however, excluding the possibility that N-terminus variants that do not affect protein stability may modify, disrupt, or attenuate other SMARCC2 functions. A clustering in this domain was not observed, thus the remaining N-terminal variants were classified as VUS due to lack of evidence. Since the analysis confirmed a LOF effect for p.(Pro77Leu), the carriers were included in the LGD group for genotype-phenotype analysis.

3D model analysis of SMARCC2 interaction with other BAF subunits indicated impairment due to missense variants in SWIRM, SANT, and CAR domains, but this could not be confirmed experimentally. Intriguingly, all mutants showed a tendency for increased interaction with SMARCC1, which, although not significant, could indicate increased formation of SMARCC1/SMARCC2 heteroduplexes or altered BAF complex assembly dynamics. No effect was seen on SMARCC2 protein localization or stability. These findings suggest that alterations in these domains have a more complex molecular pathomechanism, which is consistent with the extreme compositional complexity of the BAF complex, with over 1400 possible combinations. ^25^ Further research is needed to determine the molecular underpinnings and pathomechanisms of missense variants in these regions.

NDD with cognitive impairment, speech and motor deficits, behavioral disorders, muscular hypotonia, brain malformations, feeding difficulties or failure to thrive, short stature, and skeletal anomalies were the main clinical manifestations of the *SMARCC2*-related BAFopathy (Table 1 and 2). Facial dysmorphisms included a prominent forehead, thick eyebrows, broad/short philtrum, thin upper and thick lower lip vermilion, and outer ear malformations. Despite certain similarities to CSS and NCBRS, ^8^ the overall clinical and phenotypic manifestations of *SMARCC2* individuals appear to be non-recognizable and phenotypic hallmarks of CSS and NCBRS, including finger-/toenail hypoplasia or absence of 5th finger distal phalanges and prominent interphalangeal joints, respectively, were either absent or very rare. Less than 20% of *SMARCC2* subjects had other frequent findings like microcephaly, sparse/thin scalp hair, and hypertrichosis. The characteristic CSS brain anomaly, agenesis/dysgenesis of the corpus callosum, was also only found in a small subset of this cohort (8/34) (Table S1 and File S2 sheet “clinical_table”). In summary, the *SMARCC2*-associated phenotype has only minor resemblance to CSS and NCBRS, thus challenging the current classification as CSS8 in OMIM. Moreover, such limited resemblance complicates clinicians and geneticists to clinically suspect this BAFopathy without genetic testing.

The large number of causative variants identified in this study allowed the identification of two clinical patterns associated with truncating and non-truncating variants, respectively. Carriers of missense/in-frame variants had a more severe phenotype, especially in neurodevelopmental (Table 1), growth parameters (Table 2), and facial dysmorphisms (Figure 2A and 2B). The *de novo* occurrence of the vast majority of non-truncating variants in this cohort corroborates their severe effect. On the contrary, the impact of LGD variants were milder, explaining their frequent inheritance from a phenotypically healthy parent and their possible presence in public databases (compare p-MvT and OR-MvT in Tables 1 and 2). This association is further confirmed by the fact that the almost complete SMARCC2 protein loss, attributed to the missense variant c.230C>T, p.(Pro77Leu), was associated with milder clinical symptoms in subjects Ind-7, Ind-8, Ind-43. Overall, these results support an incomplete penetrance of loss-of-function *SMARCC2* variants, which is also found in other rare monogenic developmental disorders. ^33^

Three affected individuals (Ind-25, Ind-33, Ind-34) with *SMARCC2* LGD variants deviated from the expected clinical phenotypes. Their severe clinical presentations can be explained by a second pathogenic variant identified in other NDD-related genes. Our study suggests that loss of SMARCC2 alone cannot explain a severe phenotype. Thus, clinicians should be alerted that a severely affected individual with a *SMARCC2* LGD alteration requires further analysis for a possible second molecular diagnosis.

The two distinct neurodevelopmental presentations of *SMARCC2*-related disease suggest that missense/in-frame variants not affecting protein stability may follow a pathomechanism other than loss-of-function. One possibility is that such variants inhibit or attenuate interactions between SMARCC2 and other BAF subunits or SMARCC2 targets. Due to their clustering in evolutionarily conserved regions, these variants may exert a dominant-negative, gain-of-function or change-of-function effect. A similar mechanism has been proposed or shown for non-truncating variants in other BAF-subunits such as SMARCA4/A2, SMARCB1/E1, and DPF2. ^6, 34^

This study demonstrates that large cohorts are essential for improved characterization, standardized ascertainment of disease-associated variants and genotype-phenotype correlations in genetic diseases. *SMARCC2* clustering hotspots and recurrent variants allowed the reclassification of newly and previously reported variants, improving genetic diagnostics. Functional studies showed that N-terminal missense variants can destabilize SMARCC2 protein, although further research is needed to identify the pathomechanism for variants in other domains. Overall, individuals with *SMARCC2-*NDD exhibit non-specific clinical manifestations and lack the defining clinical characteristics of CSS and NBRS, thus requiring genetic testing for identification. By systematically analyzing and reviewing clinical data, two distinct *SMARCC2*-associated phenotypes were found: a more severe phenotype due to *de novo* non-truncating variants, and a milder phenotype due to predominately inherited LGD variants with possibly incomplete penetrance. In view of such a sharp contrast, the appropriate nomenclature allowing to distinguish the two associated clinical entities remains to be determined. The presented findings also support two distinct disease pathomechanisms underlying the corresponding clinical manifestations.

## Data Availability

The data supporting this article are provided in the supplementary files available in the online version of this article at the publisher's website or in the online repository Zenodo (Files S2, S3, S4: DOI: 10.5281/zenodo.7766791).

https://zenodo.org/record/7766791#.ZCWs-nZBwQ9

## ACKNOWLEDGMENTS

The authors thank the individuals and their families for participating in this study. Furthermore, we are indebted to GeneDX for establishing contact with the clinicians. The authors I.M., A.R., D.W., V.C., O.K., G.W.E.S., A.R. and G.V. are members of the European Reference Network on Rare Congenital Malformations and Rare Intellectual Disability ERN-ITHACA [EU Framework Partnership Agreement ID: 3HP-HP-FPA ERN-01-2016/739516]. The authors acknowledge ERN-ITHACA for posting a call for a collaborative project, which led to the recruitment of cases. Also we acknowledge TUDP for performing exome sequencing of Fam-21 with the Telethon project GSP15001.

## AUTHOR CONTRIBUTIONS

Conceptualization: A.A., G.V.; Data curation: A.A., B.P., G.V.; Formal analysis: E.B., B.P., M.S.; Investigation: E.B., E.G., B.P., G.V.; Methodology: E.B., B.P., G.V.; Project administration: G.V. Resources: M.Z., B.P., C.S., PJ. van der S., I.M., A.M.P., A.R., L.P.B., S.G., C.M., O.B., D.H., L.H., B.I., B.C., D.W., V.C., M.S., P.D.M., M.O., R.A.J., K.P., L.B.C., O.K., A. van H., R.M., I.V., I.C., JA. M-A., AM. R., HC. M., E.M.P., C.C., K. A-Y, M.W., M.C.H., P.Z., I.T., G.B., M.U., PT. B., E.M., J.M., J. M van de K., E.J.P., A.D., A.S., B.H.G., E-J.G. MK.G., F.G.R., E.AR N., G.H., C.K., G.G, R.B., J.R., MM.M., F.M., V.S., A.T., V.N., M.K., RE.S., G.W.E.S., M.Z. P.M.C., M.S., A.R., A.A., G.V. Software: N/A; Supervision: G.V.; Validation: E.B.; Visualization: E.B., B.P., A.A., G.V.; Writing-original draft: E.B., B.P., A.A, G.V.; Writing-review & editing: G.V., A.R.

## ETHICS DECLARATION

Legal guardians gave written informed consent for genetic and clinical data, including photos and brain images, to be published. This study follows the Declaration of Helsinki protocols and is approved by the ethics committees of the Friedrich-Alexander-Universität Erlangen-Nürnberg, Germany (259_16 Bc), University of Leipzig Germany (224/16-ek and 402/16-ek), Leiden University Medical Center, Leiden, The Netherlands (G21.129), Telethon Institute of Genetics and Medicine (TIGEM), Naples, Italy (number of protocol 81/21), North Ostrobothnia’s Hospital District (19.4.2021, Eettmk § 110), Greenwood Genetic Center, Self Regional Healthcare (IRB Number: Pro00085001), UMT - University of Management and Technology, Lahore, Pakistan {IRB Ref. Number: DLSBBC-2022-04}, Hospital vall d’Hebron (code C.0002416) and the Children’s Mercy Institutional Review Board (IRB) (Study # 11120514).

## SUPPLEMENTARY

**File S1 |** Supplementary notes with clinical reports, supplementary methods, results, figures and tables, references

**File S2 |** Comprehensive clinical data (DOI: 10.5281/zenodo.7766791)

**File S3 |** Comprehensive genetic data (DOI: 10.5281/zenodo.7766791)

**File S4 |** SMARCC2 N-terminal homology model (DOI: 10.5281/zenodo.7766791)

## REFERENCES

1. Staahl BT, Crabtree GR. Creating a neural specific chromatin landscape by npBAF and nBAF complexes. Curr Opin Neurobiol. 2013;23(6):903–913. doi:10.1016/j.conb.2013.09.003

2. Vasko A, Drivas TG, Schrier Vergano SA. Genotype-phenotype correlations in 208 individuals with Coffin-Siris syndrome. Genes. 2021;12(6):937. doi:10.3390/genes12060937

3. Hoyer J, Ekici AB, Endele S, et al. Haploinsufficiency of ARID1B, a member of the SWI/SNF-a chromatin-remodeling complex, is a frequent cause of intellectual disability. Am J Hum Genet. 2012;90(3):565–572. doi:10.1016/j.ajhg.2012.02.007

4. Kosho T, Okamoto N, Coffin-Siris Syndrome International Collaborators. Genotype-phenotype correlation of Coffin-Siris syndrome caused by mutations in SMARCB1, SMARCA4, SMARCE1, and ARID1A. Am J Med Genet C Semin Med Genet. 2014;166C(3):262–275. doi:10.1002/ajmg.c.31407

5. Santen GWE, Aten E, Vulto-van Silfhout AT, et al. Coffin-Siris syndrome and the BAF complex: genotype-phenotype study in 63 patients. Hum Mutat. 2013;34(11):1519–1528. doi:10.1002/humu.22394

6. Vasileiou G, Vergarajauregui S, Endele S, et al. Mutations in the BAF-complex subunit DPF2 are associated with Coffin-Siris syndrome. Am J Hum Genet. 2018;102(3):468–479. doi:10.1016/j.ajhg.2018.01.014

7. Wieczorek D, Bögershausen N, Beleggia F, et al. A comprehensive molecular study on Coffin-Siris and Nicolaides-Baraitser syndromes identifies a broad molecular and clinical spectrum converging on altered chromatin remodeling. Hum Mol Genet. 2013;22(25):5121–5135. doi:10.1093/hmg/ddt366

8. Machol K, Rousseau J, Ehresmann S, et al. Expanding the spectrum of BAF-related disorders: de novo variants in SMARCC2 cause a syndrome with intellectual disability and developmental delay. Am J Hum Genet. 2019;104(1):164–178. doi:10.1016/j.ajhg.2018.11.007

9. Li D, Downes H, Hou C, et al. Further supporting SMARCC2-related neurodevelopmental disorder through exome analysis and reanalysis in two patients. Am J Med Genet A. 2022;188(3):878–882. doi:10.1002/ajmg.a.62597

10. Chen CA, Lattier J, Zhu W, et al. Retrospective analysis of a clinical exome sequencing cohort reveals the mutational spectrum and identifies candidate disease-associated loci for BAFopathies. Genet Med Off J Am Coll Med Genet. 2022;24(2):364–373. doi:10.1016/j.gim.2021.09.017

11. Yi S, Li M, Yang Q, et al. De novo SMARCC2 variant in a chinese woman with Coffin-Siris syndrome 8: a case report with mild intellectual disability and endocrinopathy. J Mol Neurosci MN. 2022;72(6):1293–1299. doi:10.1007/s12031-022-02010-0

12. Lo T, Kushima I, Aleksic B, et al. Sequencing of selected chromatin remodelling genes reveals increased burden of rare missense variants in ASD patients from the Japanese population. Int Rev Psychiatry Abingdon Engl. 2022;34(2):154–167. doi:10.1080/09540261.2022.2072193

13. Gofin Y, Zhao X, Gerard A, et al. Evidence for an association between Coffin-Siris syndrome and congenital diaphragmatic hernia. Am J Med Genet A. 2022;188(9):2718–2723. doi:10.1002/ajmg.a.62889

14. Tuoc T, Dere E, Radyushkin K, et al. Ablation of BAF170 in developing and postnatal dentate gyrus affects neural stem cell proliferation, differentiation, and learning. Mol Neurobiol. 2017;54(6):4618–4635. doi:10.1007/s12035-016-9948-5

15. Tuoc TC, Boretius S, Sansom SN, et al. Chromatin regulation by BAF170 controls cerebral cortical size and thickness. Dev Cell. 2013;25(3):256–269. doi:10.1016/j.devcel.2013.04.005

16. Sun H, Zhang S, Wang J, et al. Expanding the phenotype associated with SMARCC2 variants: a fetus with tetralogy of Fallot. BMC Med Genomics. 2022;15(1):40. doi:10.1186/s12920-022-01185-0

17. Sobreira N, Schiettecatte F, Valle D, Hamosh A. GeneMatcher: a matching tool for connecting investigators with an interest in the same gene. Hum Mutat. 2015;36(10):928–930. doi:10.1002/humu.22844

18. Li H. Aligning sequence reads, clone sequences and assembly contigs with BWA-MEM. Published online 2013. doi:10.48550/ARXIV.1303.3997

19. DePristo MA, Banks E, Poplin R, et al. A framework for variation discovery and genotyping using next-generation DNA sequencing data. Nat Genet. 2011;43(5):491–498. doi:10.1038/ng.806

20. Lefter M, Vis JK, Vermaat M, den Dunnen JT, Taschner PEM, Laros JFJ. Mutalyzer 2: next generation HGVS nomenclature checker. Bioinforma Oxf Engl. 2021;37(18):2811–2817. doi:10.1093/bioinformatics/btab051

21. McLaren W, Gil L, Hunt SE, et al. The ensembl variant effect predictor. Genome Biol. 2016;17(1):122. doi:10.1186/s13059-016-0974-4

22. Richards S, Aziz N, Bale S, et al. Standards and guidelines for the interpretation of sequence variants: a joint consensus recommendation of the American College of Medical Genetics and Genomics and the Association for Molecular Pathology. Genet Med Off J Am Coll Med Genet. 2015;17(5):405–424. doi:10.1038/gim.2015.30

23. Köhler S, Carmody L, Vasilevsky N, et al. Expansion of the Human Phenotype Ontology (HPO) knowledge base and resources. Nucleic Acids Res. 2019;47(D1):D1018–D1027. doi:10.1093/nar/gky1105

24. Mashtalir N, Suzuki H, Farrell DP, et al. A structural model of the endogenous human BAF complex informs disease mechanisms. Cell. 2020;183(3):802–817.e24. doi:10.1016/j.cell.2020.09.051

25. He S, Wu Z, Tian Y, et al. Structure of nucleosome-bound human BAF complex. Science. 2020;367(6480):875–881. doi:10.1126/science.aaz9761

26. Allen MD, Freund SMV, Bycroft M, Zinzalla G. SWI/SNF subunit BAF155 N-terminus structure informs the impact of cancer-associated mutations and reveals a potential drug binding site. Commun Biol. 2021;4(1):528. doi:10.1038/s42003-021-02050-z

27. Kelley LA, Mezulis S, Yates CM, Wass MN, Sternberg MJE. The Phyre2 web portal for protein modeling, prediction and analysis. Nat Protoc. 2015;10(6):845–858. doi:10.1038/nprot.2015.053

28. Xi Q, He W, Zhang XHF, Le HV, Massagué J. Genome-wide impact of the BRG1 SWI/SNF chromatin remodeler on the transforming growth factor β transcriptional program. J Biol Chem. 2008;283(2):1146–1155. doi:10.1074/jbc.M707479200

29. Gregor A, Meerbrei T, Gerstner T, et al. *De novo* missense variants in FBXO11 alter its protein expression and subcellular localization. Hum Mol Genet. 2022;31(3):440–454. doi:10.1093/hmg/ddab265

30. Wittmann MT, Katada S, Sock E, et al. scRNA sequencing uncovers a TCF4-dependent transcription factor network regulating commissure development in mouse. Development. 2021;148(14):dev196022. doi:10.1242/dev.196022

31. Xiang J, Peng J, Baxter S, Peng Z. AutoPVS1: an automatic classification tool for PVS1 interpretation of null variants. Hum Mutat. 2020;41(9):1488–1498. doi:10.1002/humu.24051

32. Ittisoponpisan S, Islam SA, Khanna T, Alhuzimi E, David A, Sternberg MJE. Can predicted protein 3D structures provide reliable insights into whether missense variants are disease associated? J Mol Biol. 2019;431(11):2197–2212. doi:10.1016/j.jmb.2019.04.009

33. Kingdom R, Tuke M, Wood A, et al. Rare genetic variants in genes and loci linked to dominant monogenic developmental disorders cause milder related phenotypes in the general population. Am J Hum Genet. 2022;109(7):1308–1316. doi:10.1016/j.ajhg.2022.05.011

34. Kosho T, Miyake N, Carey JC. Coffin-Siris syndrome and related disorders involving components of the BAF (mSWI/SNF) complex: historical review and recent advances using next generation sequencing. Am J Med Genet C Semin Med Genet. 2014;166C(3):241–251. doi:10.1002/ajmg.c.31415

35. Aravind L, Iyer LM. The SWIRM domain: a conserved module found in chromosomal proteins points to novel chromatin-modifying activities. Genome Biol. 2002;3(8):research0039.1-0039.7. doi:10.1186/gb-2002-3-8-research0039

36. Da G, Lenkart J, Zhao K, Shiekhattar R, Cairns BR, Marmorstein R. Structure and function of the SWIRM domain, a conserved protein module found in chromatin regulatory complexes. Proc Natl Acad Sci. 2006;103(7):2057–2062. doi:10.1073/pnas.0510949103

37. Grüne T, Brzeski J, Eberharter A, et al. Crystal structure and functional analysis of a nucleosome recognition module of the remodeling factor ISWI. Mol Cell. 2003;12(2):449–460. doi:10.1016/S1097-2765(03)00273-9

38. Boyer LA, Latek RR, Peterson CL. The SANT domain: a unique histone-tail-binding module? Nat Rev Mol Cell Biol. 2004;5(2):158–163. doi:10.1038/nrm1314

39. Rentzsch P, Witten D, Cooper GM, Shendure J, Kircher M. CADD: predicting the deleteriousness of variants throughout the human genome. Nucleic Acids Res. 2019;47(D1):D886–D894. doi:10.1093/nar/gky1016

