## Supplementary material for "Elucidating the clinical and molecular spectrum of *SMARCC2*-associated NDD in a cohort of 65 affected individuals": File S1

### SUPPLEMENTARY METHODS

#### Cohort

Two out of 41 novel individuals were included in previous publications with either incomplete clinical or molecular findings. More specifically, the Ind-18 variant (c.172C>T p.(Gln58*)) had previously been described in a publication, but the clinical description was missing. [1] Ind-25, with the variant c.1094_1097del p.(Lys365Thrfs*12), was also reported in a recent case report and was included in the current cohort due to the description of significant additional clinical and molecular findings. [2] The novel individuals were recruited either via an international collaborative network of research and diagnostic sequencing centers or by employing the web-based matching platform GeneMatcher [3] and posting a call for a collaborative project in ERN-ITHACA (European Network of Rare Malformation Syndromes).

#### Genetic analysis

Two of the five affected individuals in Fam-18 (Ind-22 and Ind-23) and Ind-42 were identified by Sanger sequencing as having the familial *SMARCC2* variant. Targeted Sanger sequencing utilizing standard methods was also performed, either for confirmation of identified variants or segregation analysis in the absence of trio exome data.

The previously published *SMARCC2* variants of 17 clinically characterized individuals [2,4,5], five cases with DD/ID or autism spectrum disorder (ASD) without further clinical description, a fetus with cardiac defects and a newborn with a diaphragmatic hernia who died in the perinatal period [1,6–8] were reviewed and included in File S2 sheets "clinical_table" and protein linear model.

#### Clinical Information

Using a standardized questionnaire, clinical, phenotypic, and neuroimaging data were collected from the referring physicians. For the extraction of clinical information on previously reported individuals, the same questionnaire was used. Facial features were determined independently by two clinical geneticists, either from the respective clinical report or from analysis of the published pictures. Brain MRI data were reviewed by an experienced pediatric neuroradiologist. For the cases without available MRI images neuroimaging data were obtained from the reports. Face2Gene research application was applied to a single 2D frontal facial photograph of a total of 23 novel and previously published individuals (10 with missense/in-frame and 13 with LGD variants). Individuals wearing glasses in the available photos or carrying a second pathogenic/likely pathogenic variant in another gene were excluded.

**RNA isolation, RT-PCR and qRT-PCR**

Total RNA was extracted from untransformed blood lymphocytes of Ind-19 and his unaffected brother, Ind-29 and one control individual with the PAXgene Blood System (Becton Dickinson). cDNA was prepared with a Superscript II Reverse Transcriptase Kit (Invitrogen, Carlsbad, CA, USA), according to the manufacturer's instructions. RT-PCR for In-29 was performed with the Platinum PCR SuperMix High Fidelity kit (Invitrogen), using primers encompassing exons 18-20 of *SMARCC2* gene (NM_003075.5) (18F: 5'- CAGACCTCTGCTTCCCAAC-3' and 20R: 5'-CAGCAGCAGAGGCGACTC-3'). PCR products were purified using the GenUP Exo SAP kit (BiotechRabbit, Berlin, Germany) and sequenced by the Big Dye Terminator Cycle Sequencing kit (Applied Biosystems, Foster City, CA, USA). *SMARCC2* expression levels of Ind-19 were analyzed by qRT-PCR with predesigned Taqman gene-expression assay (SMARCC2: Hs00161961_m1) on an ABI 7900HT instrument (ThermoFisher Scientific). For normalization of results to the mean, the following endogenous controls were used: b-actin (huACTB), b-2 microglobulin (huB2M), acidic ribosomal protein (huPO), and phosphoglycerate kinase 1 (huPGK1) (all Thermo Fisher Scientific). Reactions were performed in four replicates. mRNA levels were calculated with the DDCt method.

**Cell Culture**

HEK293T cells were grown in Dulbecco's Modified Eagle's Medium (DMEM) supplemented with 10% fetal calf serum (FCS) and 1% penicillin/streptomycin at 37°C in 5% CO2. Plasmids were transfected with JetPrime® (Polyplus Life Science) according to manufacturer's instructions.

**Plasmids and Mutagenesis**

Flag-tagged SMARCC2 was obtained from addgene (plasmid #19142) and has previously been described [9]. Mutagenesis was carried out using the In-Fusion HD Cloning kit (Clontech). Oligonucleotide sequences are provided in Table S4.

**Immunofluorescence, PLA and Microscopy**

Cells were grown on coverslips. 24 h after transfection, they were fixed in 3.7 % formaldehyde for 10 min at room temperature, permeabilized for 10 min in 0.5 % Triton-X-100 and washed in PBS. Antibodies were diluted in PBS containing 5 % normal goat serum. Cells were incubated with the primary antibody (mouse monoclonal FLAG, Sigma F1804) for 1 h at 37 °C and washed in PBS containing 0.1 % Tween20, followed by incubation with the Cy3-conjugated secondary antibody (dianova 115-165-146) for 1 h at 37 °C and another wash. Coverslips were mounted using ProLong™ Antifade mountant (Thermo Fisher Scientific).

For PLA, cells were seeded onto coverslips 17 h post transfection and fixed and permeabilized as described above 30 h post transfection. PLA was performed using Duolink In Situ reagents (Sigma) according to the manufacturer's instructions. All slides were imaged on a Zeiss AxioImager Z2 with Apotome using the AxioVision software and processed using AxioVision and ImageJ.

#### Immunoprecipitation and Western Blotting

Whole cell lysate was generated as previously described. [10] Briefly, cells were washed in PBS and rotated in buffer A (10 mM HEPES, pH 7.9; 10 mM KCl; 0.1 mM EDTA, pH 8.0; 0.1 mM EGTA, pH 8.0; 2 mM DTT; 10 µg/ml Aprotinin; 10 µg/ml Leupeptin; 1 % NP-40; 300 mM NaCl) for 15 min at 4 °C. Following centrifugation at 16000 x g for 5 min, protein concentration was determined using the Qubit™ Protein Assay-kit (Thermo Fisher Scientific).

For Co-immunoprecipitation of FLAG-tagged SMARCC2, 2 mg whole cell lysate of transfected or untransfected cells (to control for nonspecific antibody binding) was incubated in CoIP-buffer (10 mM HEPES, pH 7.9; 0.1 mM EDTA, 0.1 mM EGTA, 120 mM NaCl, 10 % glycerol) at 4 °C overnight with 4 µg of mouse monoclonal FLAG (Sigma F1804), 8 µg of rabbit polyclonal FLAG (Sigma F7425) or no antibody (to control for nonspecific binding to beads) and pulled down by incubation for 3 h at 4 °C with magnetic SureBeads™ (BioRad). Following 3 washes in CoIP-buffer, proteins were eluted twice in 50 µl 2x Lämmli-buffer at 90 °C for 5 minutes each.

15 µl of the IP fraction were run on 4-15 % gradient polyacrylamide gels and transferred onto nitrocellulose membranes using semi-dry blotting. Membranes were blocked in 5 % non-fat dry milk in TBS and incubated with antibodies in 3 % non-fat dry milk in TBS-T. Incubation with primary antibodies occurred overnight at 4 °C. Following three washes in TBS-T, secondary antibodies were incubated for 1 h at room temperature. Membranes were developed with SuperSignal™ West Femto Maximum Sensitivity Substrate (Thermo Scientific) and imaged on a ChemiDoc Imaging System (BioRad). Protein bands were quantified by densitometry using the software ImageLab (BioRad).

#### Antibodies

All antibodies used in this work are listed in Table S5.

### SUPPLEMENTARY RESULTS

#### Additional variants in SMARCC2 individuals

Along with the *SMARCC2* frameshift variant, Ind-25 and Ind-33 carried a likely pathogenic splice variant in the *TANC2* gene (MIM 615047), which is linked to an NDD with autistic features and language delay (MIM 618906) and a pathogenic 1p36 microdeletion, respectively. In addition to the *de novo* in-frame *SMARCC2* variant, Ind-34 had a *de novo* likely pathogenic missense substitution in the *ACSL4* gene, which was linked to the NDD X-linked 63 entity (MIM 300387). Of note, Ind-42 additionally carried a maternally inherited pathogenic *RAF1* (MIM 164760) variant linked to dilated cardiomyopathy (MIM 615916); this was not detected in the affected older sister (Ind-35).

#### Prevalence of clinical manifestations in literature vs novel *SMARCC2* cases

The majority of the clinical manifestations and dysmorphic facial features were found to have similar frequencies between the previously published *SMARCC2* subjects and the novel cohort (Table 1 and 2 and Table S1). Moderate/severe global DD/ID and muscular hypotonia seemed to be more common in the literature, while mild developmental impairment and autistic behavior described more often in this cohort. The novel cohort had a lower prevalence for both congenital anomalies of the genitourinary system and gastrointestinal tract as well as for the facial feature broad philtrum. However, after correction for multiple testing, p-values for all aforementioned traits remained not significant. The only clinical features which remained significantly lower in novel subjects after correction were the eye structural abnormalities and outer ear anomalies (FDR corrected p-value: 0.029 and 0.034, respectively) (Table 2).

#### Clinical reports

##### Ind-01 case report

Removed to comply with medRxiv policy.

##### Ind-02 case report

Removed to comply with medRxiv policy.

##### Ind-03 case report

Removed to comply with medRxiv policy.

##### Ind-04 case report

Removed to comply with medRxiv policy.

**Ind-05 case report**

Removed to comply with medRxiv policy.

**Ind-06 case report**

Removed to comply with medRxiv policy.

**Ind-07 case report**

Removed to comply with medRxiv policy.

**Ind-08 case report**

Removed to comply with medRxiv policy.

**Ind-09 case report**

Removed to comply with medRxiv policy.

**Ind-10 case report**

Removed to comply with medRxiv policy.

**Ind-11 case report**

Removed to comply with medRxiv policy.

**Ind-12 case report**

Removed to comply with medRxiv policy.

**Ind-13 case report**

Removed to comply with medRxiv policy.

**Ind-14 case report**

Removed to comply with medRxiv policy.

**Ind-15 case report**

Removed to comply with medRxiv policy.

**Ind-16 (Family-16) case report**

Removed to comply with medRxiv policy.

**Ind-17 (Family-16) case report**

Removed to comply with medRxiv policy.

**Ind-18 case report**

Removed to comply with medRxiv policy.

**Family 18 (Ind-19, Ind-20, Ind-21, Ind-22, Ind-23)**

Removed to comply with medRxiv policy.

**Ind-19 case report**

Removed to comply with medRxiv policy.

**Ind-20 case report**

Removed to comply with medRxiv policy.

**Ind-21 case report**

Removed to comply with medRxiv policy.

**Ind-22 case report**

Removed to comply with medRxiv policy.

**Ind-23 case report**

Removed to comply with medRxiv policy.

**Ind-25 case report**

Removed to comply with medRxiv policy.

**Ind-26 case report**

Removed to comply with medRxiv policy.

**Ind-29 case report**

Removed to comply with medRxiv policy.

**Ind-31 case report**

Removed to comply with medRxiv policy.

**Ind-32 case report**

Removed to comply with medRxiv policy.

**Ind-33 case report**

Removed to comply with medRxiv policy.

**Ind-34 case report**

Removed to comply with medRxiv policy.

**Ind-35 (Family-26) case report**

Removed to comply with medRxiv policy.

**Ind-36 case report**

Removed to comply with medRxiv policy.

**Ind-37 case report**

Removed to comply with medRxiv policy.

**Ind-38 case report**

Removed to comply with medRxiv policy.

**Ind-39 case report**

Removed to comply with medRxiv policy.

**Ind-40 case report**

Removed to comply with medRxiv policy.

**Ind-41 case report**

Removed to comply with medRxiv policy.

**Ind-42 (Family-26) case report**

Removed to comply with medRxiv policy.

**Ind-43 case report**

Removed to comply with medRxiv policy.

**Ind-45 case report**

Removed to comply with medRxiv policy.

### SUPPLEMENTARY FIGURES

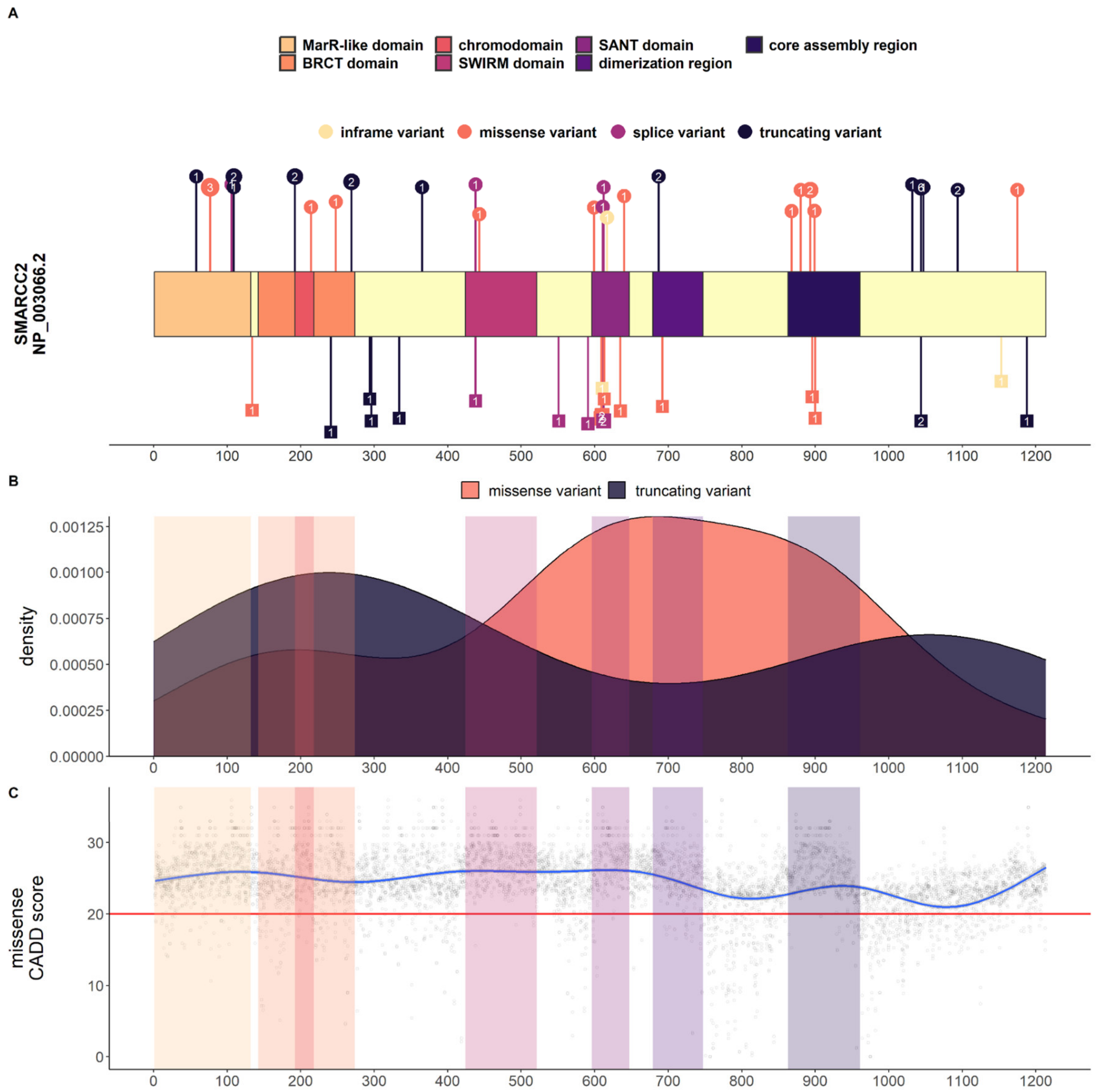


**Figure S1 | SMARCC2 domain structure with variant distribution, sequence conservation and clustering regions**

**(A)** Linear model of SMARCC2 (see main Figure 1A for variant descriptions). **(B)** Density plot depicting distribution of missense and truncating variants over the protein length. Missense variants are found mainly in the central region of the protein, occurring in the SANT, DR and CAR domains, but a cluster is also present in the N-terminal module. Truncating variants instead are found throughout the protein. **(C)** Generalized linear model of CADD PHRED scores for all possible missense variants of *SMARCC2*. The blue line shows smoothed CADD values, while the red horizontal line shows the recommended score cutoff (20). Shaded areas indicate protein domains. SMARCC2 and its functional domains are highly conserved.

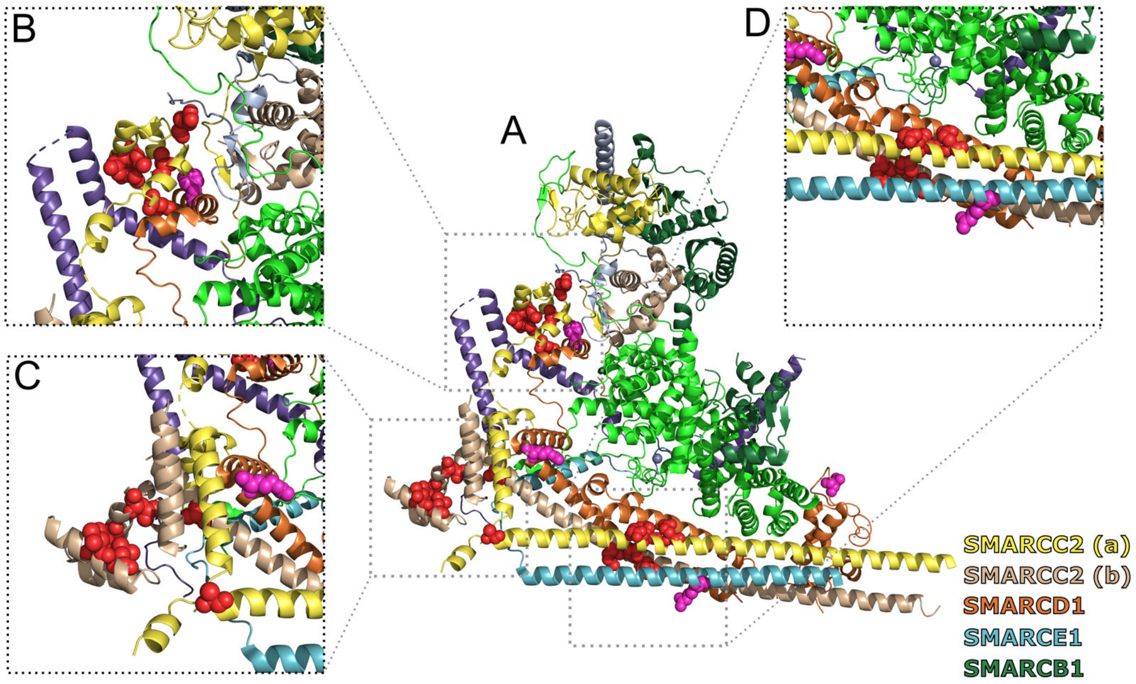


**Figure S2 | Non-truncating variants in SMARCC2 in the BAF 3D-model by He *et al.***

**(A)** Cartoon model of the BAF complex (based on PDB structure 6LTH [11]). For experimental reasons, this model was built on a BAF complex containing not a SMARCC1/SMARCC2 heterodimer, but a SMARCC2/SMARCC2 homodimer. The two chains are named SMARCC2 (a) and SMARCC2 (b) respectively. Because the SMARCC1/C2 BAF complex generated an almost identical cryo-EM map and both SMARCC proteins contain the same highly conserved domains and bind other BAF subunits in a similar manner [11], the assumptions made on the CC2/CC2 model are transferable to the CC1/CC2 model. The subunits contained in this model are indicated. The herein described novel variants are depicted in red, previously published variants in magenta. **(B)** and **(C)** Zoom of the SANT domains of the respective SMARCC2 (a) and (b) proteins. The non-truncating variants affecting amino acid positions Arg599, Leu609, Leu610, Leu613, Cys635, and Leu640 are depicted as red spheres. Note the close proximity of two of the only three described SMARCD1 missense variants [12] indicated in magenta: c.1483T>C p.(Phe495Leu) in (B) and c.1336A>G p.(Arg446Gly) in (C), respectively. The regions affected by these variants form the palm and thumb of the base module scaffold as defined by He and colleagues [11]. The close proximity of missense variants affecting these regions makes it seem likely that they disrupt the organization of the base scaffold. **(D)** Zoom of the coiled-coiled domains of the SMARCC2 proteins. The herein described novel cluster of SMARCC2 missense variants is depicted as red spheres. The affected amino acids (Ala868, Glu893, Met896) are in very close proximity to each other and also to their counterparts in the second SMARCC unit. One of the only five published SMARCE1 (NM_003079.4) missense variants (c.752G>A, p.(Arg251Gln), depicted in magenta) [13] is localized in the zinc finger interacting with the SMARCC fingers. This observation again pinpoints that pathogenic amino acid changes in this region could cause alterations in the subunit interactions and dynamics.

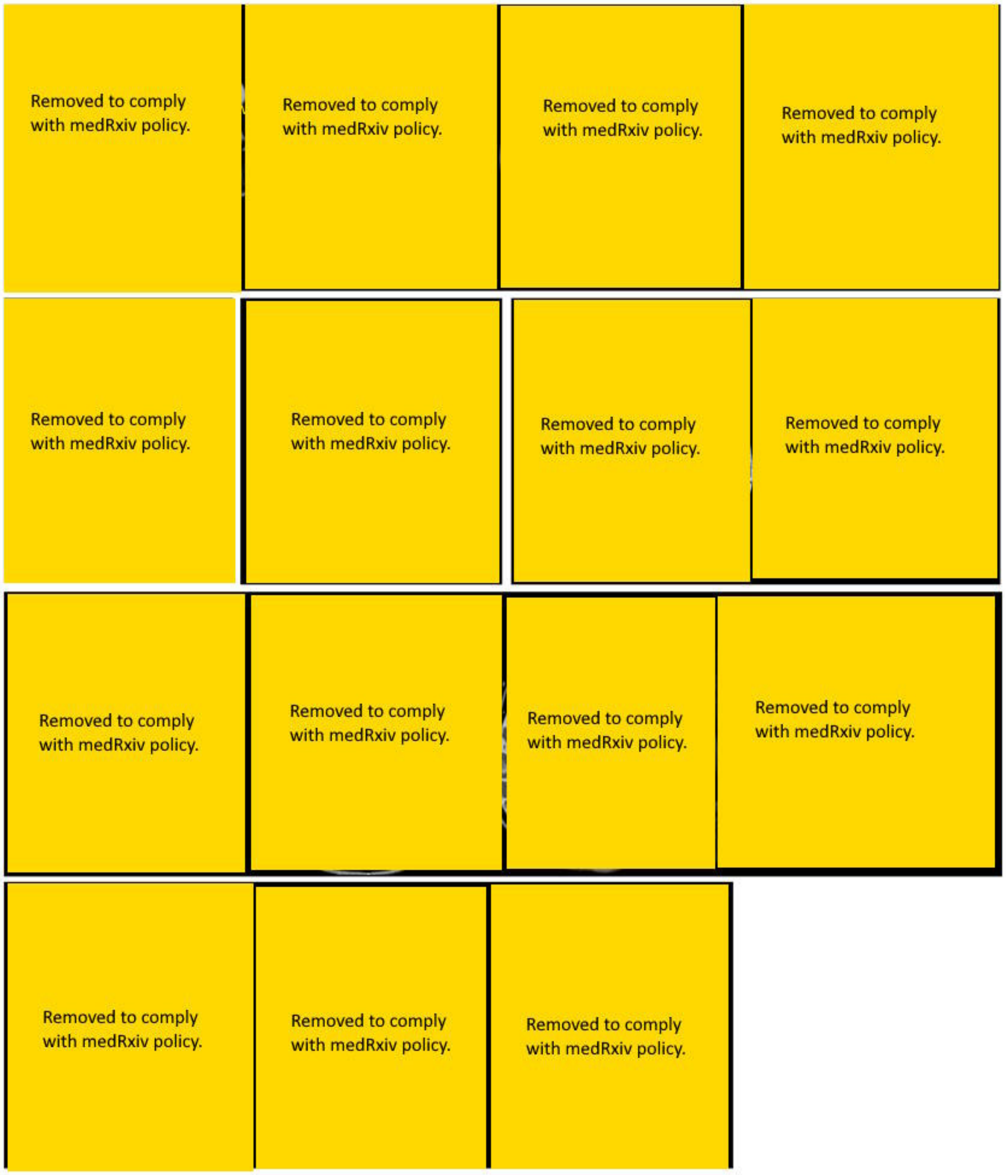


**Figure S3 | White matter involvement and additional neuroimaging findings in individuals with SMARCC2 variants**

Multiple non-specific white matter signal alterations are noted both in the parietal and frontal periventricular regions (arrowheads) variably associated with white matter volume loss and enlargement of the lateral ventricles (asterisks) and enlargement of the cerebral CSF spaces (empty arrows). Arachnoid cysts in the left temporal region and posterior cranial fossa are noted respectively in Ind-2 and Ind-11 (thin arrows). Partial ectopic neurohypophysis with T1 hyperintensity extending from the posterior lobe along the pituitary stalk is noted in Ind-25 (dotted arrow). Multiple chronic arterial ischemic infarcts in the left temporal lobe and thalamus (thick arrows) are present in Ind-33 with a normal arterial MR angiography, suggesting possible cerebral embolism.

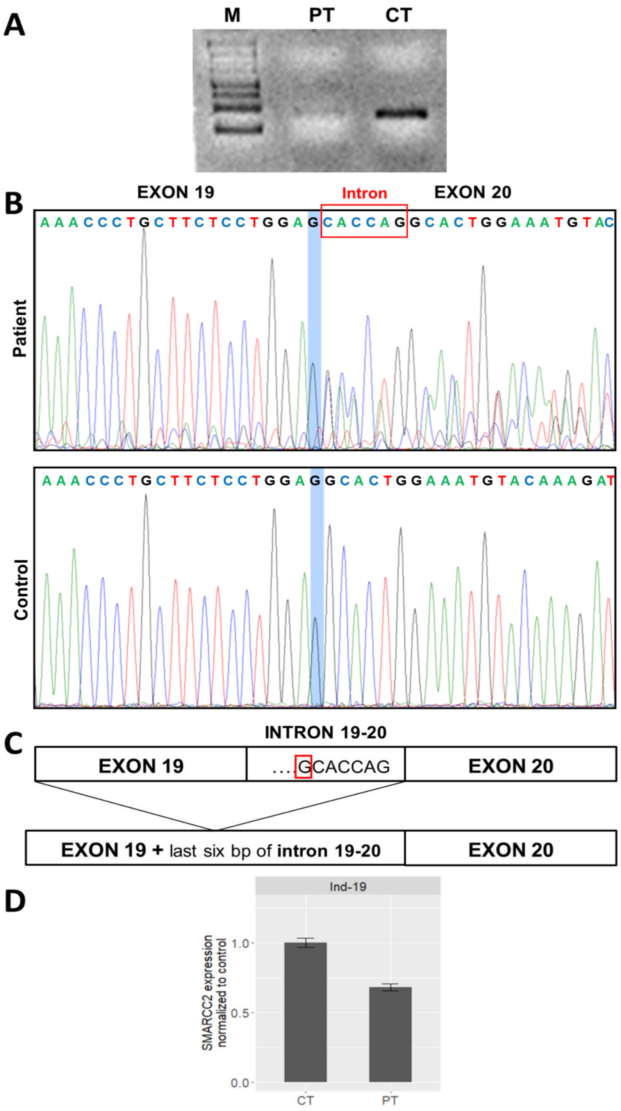


**Figure S4 | Expression assays for *SMARCC2* variants c.1834-7C>G and c.3129del**

**(A)** Agarose gel electrophoresis of RT-PCR products of Ind-29 (PT) and control individual (CT) amplified using total RNA extracted from peripheral blood. Detection of residual aberrant product in the individual carrying the c.1834-7C>G variant. M: 100 bps ladder. **(B)** Sanger sequencing of RT-PCR products. Blue stripe indicates the breakpoint of junction between the exon 19 and the last six bps of intron 19, followed by exon 20. **(C )** Schematic representation of the alternative splicing of *SMARCC2*. **(D)** qRT-PCR for *SMARCC2* in Ind-19 (PT) with the variant c.3129del p.(Gly1044Aspfs*17) showing NMD with 68% residual *SMARCC2* RNA expression. One non-carrier sibling was used as control.

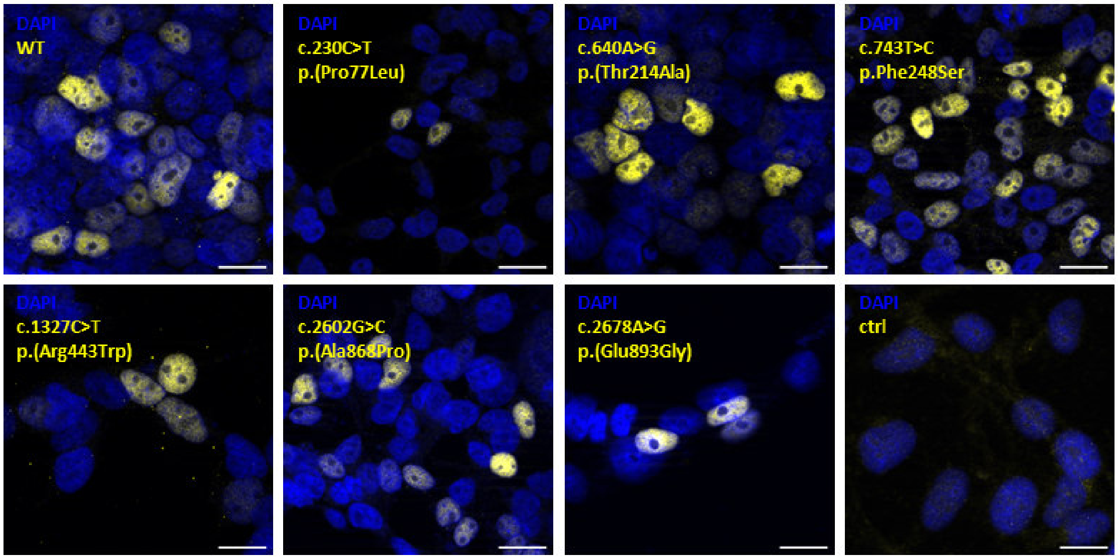


**Figure S5 | Functional analysis of protein localization**

Immunofluorescence staining of transiently overexpressed FLAG-tagged SMARCC2 protein and mutants in HEK293T cells. Yellow: FLAG; blue: DAPI; scale bar: 20 µm. The observed nuclear expression pattern is unchanged in the mutants.

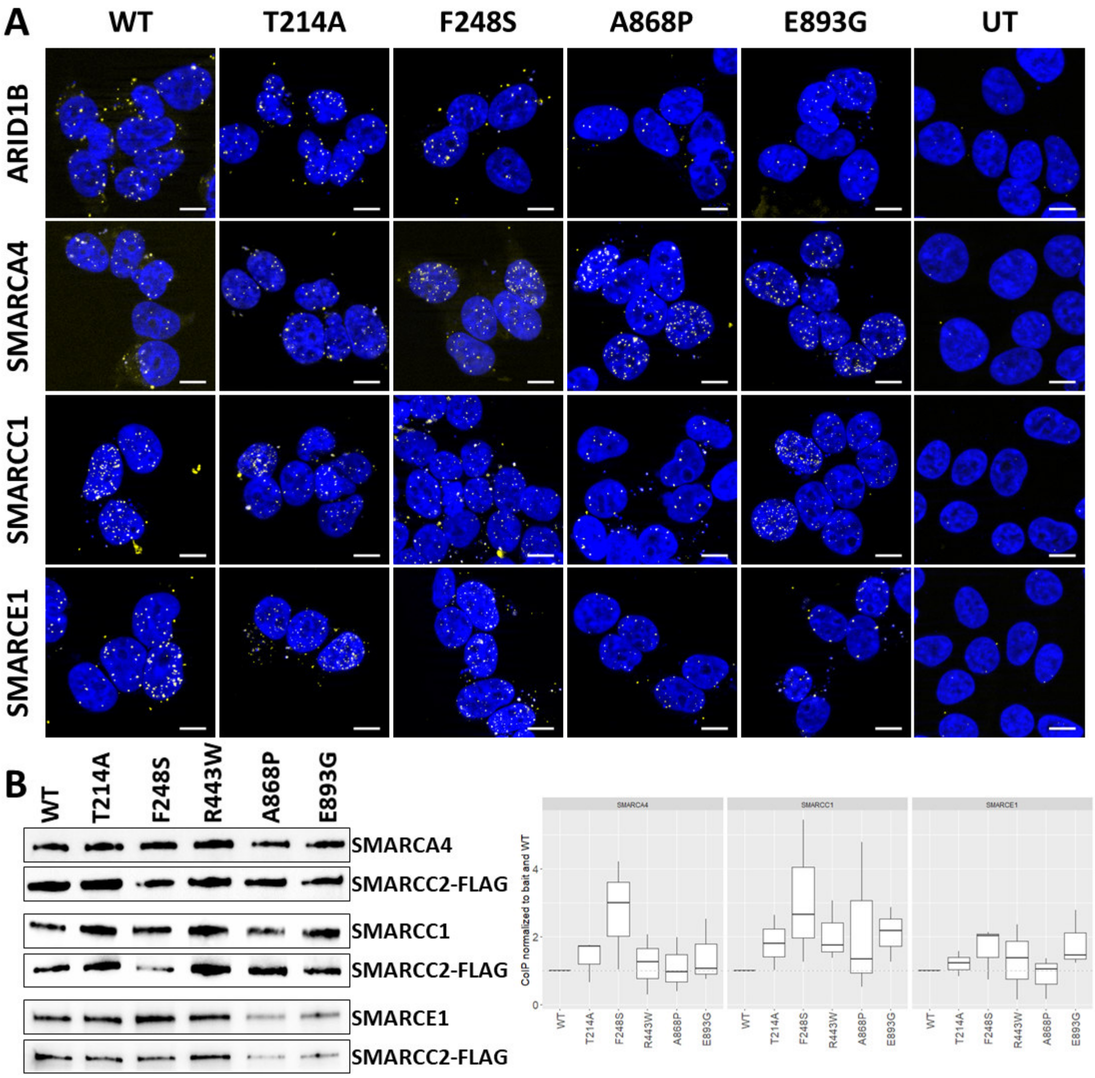


**Figure S6 | Functional analysis of protein interaction**

**(A)** Proximity ligation assay (PLA) of HEK293T cells with transiently overexpressed wildtype and mutant FLAG-tagged SMARCC2. Detection of interaction of FLAG-SMARCC2 with the BAF subunits ARID1B, SMARCA4, SMARCC1, SMARCE1. UT: untransfected; yellow: positive PLA signal indicating interaction; blue: DAPI; scale bar: 10 µm. **(B)** Co-Immunoprecipitation (CoIP) of FLAG-SMARCC2. Left panel: representative western blot images of endogenous target proteins (SMARCA4, SMARCC1, SMARCE1) and the corresponding bait signal (SMARCC2-FLAG). Right panel: box plots of signal quantification normalized to the bait protein (SMARCC2-FLAG) and wildtype sample. Data generated from three independent experiments. P-values were calculated using a one sample t-test (hypothetical mean = 1, significance threshold < 0.05). No significant changes in interactions were detected for all mutants. The interaction of all investigated SMARCC2 mutants with SMARCC1 is increased, but the effect is not statistically significant.

### SUPPLEMENTARY TABLES

#### Table S1 | Overview of all clinical characteristics of SMARCC2 individuals

| Group | Phenotype | HPO | Novel cases | Literature cases | Truncating variant | Missense/inframe variant | All cases |
|---|---|---|---|---|---|---|---|
| Intellectual and social development | Global developmental delay; Intellectual disability | HP:0001263; HP:0001249 | 82%, (31/38) | 94%, (16/17) | 72%, (21/29) | 100%, (26/26) | 85%, (47/55) |
| | mild GDD/ID | HP:0011342; HP:0001256 | 42%, (16/38) | 29%, (5/17) | 55%, (16/29) | 19%, (5/26) | 38%, (21/55) |
| | moderate/severe GDD/ID | HP:0011343; HP:0002342; HP:0011344; HP:0010864 | 39%, (15/38) | 65%, (11/17) | 17%, (5/29) | 81%, (21/26) | 47%, (26/55) |
| | Autistic behavior | HP:0000729 | 42%, (16/38) | 17%, (3/18) | 35%, (11/31) | 32%, (8/25) | 34%, (19/56) |
| | Behavioral abnormalities | HP:0000708 | 61%, (23/38) | 60%, (9/15) | 62%, (18/29) | 58%, (14/24) | 60%, (32/53) |
| Neurological system | Muscular hypotonia | HP:0001252 | 61%, (23/38) | 88%, (15/17) | 52%, (15/29) | 88%, (23/26) | 69%, (38/55) |
| | Brain imaging abnormality | HP:0410263 | 60%, (12/20) | 62%, (8/13) | 50%, (7/14) | 68%, (13/19) | 61%, (20/33) |
| | Visual impairment | HP:0000505 | 32%, (12/38) | 36%, (5/14) | 21%, (6/29) | 48%, (11/23) | 33%, (17/52) |
| | Seizures | HP:0001250 | 27%, (10/37) | 29%, (5/17) | 25%, (7/28) | 31%, (8/26) | 28%, (15/54) |
| | EEG abnormality | HP:0002353 | 21%, (6/29) | 100%, (1/1) | 20%, (4/20) | 30%, (3/10) | 23%, (7/30) |
| | Muscular hypertonia | HP:0001276 | 18%, (7/38) | 25%, (4/16) | 17%, (5/29) | 24%, (6/25) | 20%, (11/54) |
| | Hearing impairment | HP:0000365 | 11%, (4/38) | 13%, (2/15) | 10%, (3/29) | 12%, (3/24) | 11%, (6/53) |
| Craniofacial anomalies | Abnormality of the outer ear | HP:0000356 | 35%, (13/37) | 100%, (7/7) | 30%, (8/27) | 71%, (12/17) | 45%, (20/44) |
| | Thin upper lip vermilion | HP:0000219 | 41%, (15/37) | 50%, (8/16) | 48%, (14/29) | 38%, (9/24) | 43%, (23/53) |
| | Thick eyebrows | HP:0000574 | 35%, (13/37) | 44%, (7/16) | 38%, (11/29) | 38%, (9/24) | 38%, (20/53) |
| | Broad philtrum | HP:0000289 | 22%, (8/37) | 70%, (7/10) | 28%, (8/29) | 39%, (7/18) | 32%, (15/47) |
| | Prominent forehead | HP:0011220 | 30%, (11/37) | 43%, (3/7) | 26%, (7/27) | 41%, (7/17) | 32%, (14/44) |
| | Thick lower lip vermilion | HP:0000179 | 22%, (8/37) | 50%, (8/16) | 24%, (7/29) | 38%, (9/24) | 30%, (16/53) |
| | Short philtrum | HP:0000322 | 27%, (10/37) | 36%, (4/11) | 28%, (8/29) | 32%, (6/19) | 29%, (14/48) |
| | Long eyelashes | HP:0000527 | 22%, (8/37) | 47%, (7/15) | 21%, (6/29) | 39%, (9/23) | 29%, (15/52) |
| | Thick alae nasi | HP:0009928 | 24%, (9/37) | 36%, (5/14) | 24%, (7/29) | 32%, (7/22) | 27%, (14/51) |
| | Wide nose | HP:0000445 | 22%, (8/37) | 36%, (5/14) | 24%, (7/29) | 27%, (6/22) | 25%, (13/51) |
| | Down-slanting palpebral fissures | HP:0000494 | 16%, (6/37) | 36%, (4/11) | 21%, (6/29) | 21%, (4/19) | 21%, (10/48) |
| | Upturned nasal tip | HP:0000463 | 14%, (5/37) | 27%, (4/15) | 10%, (3/29) | 26%, (6/23) | 17%, (9/52) |
| | Wide mouth | HP:0000154 | 14%, (5/37) | 20%, (2/10) | 7%, (2/29) | 28%, (5/18) | 15%, (7/47) |
| | Hypertelorism | HP:0000316 | 11%, (4/37) | 25%, (3/12) | 10%, (3/29) | 20%, (4/20) | 14%, (7/49) |
| | Depressed nasal bridge | HP:0005280 | 11%, (4/37) | 14%, (2/14) | 10%, (3/29) | 14%, (3/22) | 12%, (6/51) |

| Group | Phenotype | HPO | Novel cases | Literature cases | Truncating variant | Missense/inframe variant | All cases |
|---|---|---|---|---|---|---|---|
| | Macrotia | HP:0000400 | 5%, (2/37) | 11%, (1/9) | 7%, (2/28) | 6%, (1/18) | 7%, (3/46) |
| | Epicanthic fold | HP:0000286 | 5%, (2/37) | 9%, (1/11) | 3%, (1/29) | 11%, (2/19) | 6%, (3/48) |
| | Up-slanting palpebral fissures | HP:0000582 | 5%, (2/37) | 0%, (0/11) | 0%, (0/29) | 11%, (2/19) | 4%, (2/48) |
| | Downturned mouth | HP:0002714 | 3%, (1/37) | 0%, (0/11) | 3%, (1/29) | 0%, (0/19) | 2%, (1/48) |
| Phenotypical abnormalities of body and face | Abnormal facial shape | HP:0001999 | 46%, (17/37) | 43%, (6/14) | 36%, (10/28) | 57%, (13/23) | 45%, (23/51) |
| | Abnormality of skeletal morphology | HP:0011842 | 31%, (11/35) | 38%, (6/16) | 26%, (7/27) | 42%, (10/24) | 33%, (17/51) |
| | Abnormality of the hand | HP:0001155 | 33%, (12/36) | 25%, (4/16) | 29%, (8/28) | 33%, (8/24) | 31%, (16/52) |
| | Decreased body weight | HP:0004325 | 26%, (9/35) | 40%, (6/15) | 11%, (3/27) | 52%, (12/23) | 30%, (15/50) |
| | Abnormality of the foot | HP:0001760 | 29%, (10/35) | 31%, (5/16) | 26%, (7/27) | 33%, (8/24) | 29%, (15/51) |
| | Abnormality of the eye | HP:0000478 | 16%, (6/37) | 73%, (8/11) | 8%, (2/26) | 55%, (12/22) | 29%, (14/48) |
| | Short stature | HP:0004322 | 20%, (7/35) | 33%, (5/15) | 7%, (2/27) | 43%, (10/23) | 24%, (12/50) |
| | Macrocephaly | HP:0000256 | 15%, (5/34) | 21%, (3/14) | 16%, (4/25) | 17%, (4/23) | 17%, (8/48) |
| | Microcephaly | HP:0000252 | 12%, (4/34) | 7%, (1/14) | 4%, (1/25) | 17%, (4/23) | 10%, (5/48) |
| Skeletal anomalies | Scoliosis | HP:0002650 | 26%, (10/38) | 31%, (5/16) | 17%, (5/29) | 40%, (10/25) | 28%, (15/54) |
| | Joint laxity | HP:0001388 | 16%, (6/37) | 7%, (1/14) | 21%, (6/28) | 4%, (1/23) | 14%, (7/51) |
| | Delayed skeletal maturation | HP:0002750 | 16%, (3/19) | 0%, (0/14) | 6%, (1/16) | 12%, (2/17) | 9%, (3/33) |
| | Clinodactyly | HP:0030084 | 8%, (3/37) | 7%, (1/15) | 7%, (2/29) | 9%, (2/23) | 8%, (4/52) |
| | Brachydactyly | HP:0001156 | 5%, (2/37) | 7%, (1/15) | 3%, (1/29) | 9%, (2/23) | 6%, (3/52) |
| | Prominent interphalangeal joints | HP:0006237 | 3%, (1/35) | 6%, (1/16) | 4%, (1/27) | 4%, (1/24) | 4%, (2/51) |
| | Pectus excavatum | HP:0000767 | 3%, (1/38) | 7%, (1/14) | 0%, (0/29) | 9%, (2/23) | 4%, (2/52) |
| | Craniosynostosis | HP:0001363 | 3%, (1/37) | 0%, (0/14) | 4%, (1/28) | 0%, (0/23) | 2%, (1/51) |
| | Absent distal phalanges of 5th finger | HP:0005807 | 0%, (0/36) | 0%, (0/16) | 0%, (0/28) | 0%, (0/24) | 0%, (0/52) |
| Ectodermal anomalies | Hypertrichosis | HP:0000998 | 8%, (3/38) | 38%, (6/16) | 14%, (4/29) | 20%, (5/25) | 17%, (9/54) |
| | Sparse/thin scalp hair | HP:0002209 | 11%, (4/38) | 29%, (5/17) | 7%, (2/29) | 27%, (7/26) | 16%, (9/55) |
| | Delayed eruption of primary teeth | HP:0000680 | 0%, (0/21) | 100%, (2/2) | 0%, (0/15) | 25%, (2/8) | 9%, (2/23) |
| | Delayed eruption of permanent teeth | HP:0000696 | 0%, (0/18) | 100%, (1/1) | 0%, (0/12) | 14%, (1/7) | 5%, (1/19) |
| | Aplasia/Hypoplasia of the nails | HP:0008386 | 3%, (1/37) | 7%, (1/15) | 0%, (0/29) | 9%, (2/23) | 4%, (2/52) |
| | Microdontia | HP:0000691 | 0%, (0/34) | NaN%, (0/0) | 0%, (0/25) | 0%, (0/9) | 0%, (0/34) |
| Congenital anomalies | Abnormality of the gastrointestinal tract | HP:0011024 | 17%, (6/36) | 100%, (3/3) | 12%, (3/24) | 40%, (6/15) | 23%, (9/39) |
| | Abnormality of the genitourinary system | HP:0000119 | 17%, (6/35) | 100%, (2/2) | 13%, (3/23) | 36%, (5/14) | 22%, (8/37) |
| | Generalized abnormality of skin | HP:0011354 | 8%, (3/38) | 35%, (6/17) | 10%, (3/29) | 23%, (6/26) | 16%, (9/55) |
| | Abnormal heart morphology | HP:0001627 | 8%, (3/36) | 23%, (3/13) | 11%, (3/28) | 14%, (3/21) | 12%, (6/49) |

| Group | Phenotype | HPO | Novel cases | Literature cases | Truncating variant | Missense/inframe variant | All cases |
|---|---|---|---|---|---|---|---|
| | Hernia | HP:0100790 | 8%, (3/36) | 20%, (3/15) | 4%, (1/27) | 21%, (5/24) | 12%, (6/51) |
| | Abnormality of the kidney | HP:0000077 | 9%, (3/34) | NaN%, (0/0) | 5%, (1/22) | 17%, (2/12) | 9%, (3/34) |
| | Laryngotracheomalacia | HP:0008755 | 3%, (1/37) | 13%, (2/15) | 4%, (1/28) | 8%, (2/24) | 6%, (3/52) |
| Miscellaneous | Feeding difficulties/failure to thrive | HP:0011968; HP:0001508 | 47%, (17/36) | 59%, (10/17) | 33%, (9/27) | 69%, (18/26) | 51%, (27/53) |
| | Sleep disturbance | HP:0002360 | 22%, (8/37) | 20%, (3/15) | 21%, (6/28) | 21%, (5/24) | 21%, (11/52) |
| | Recurrent infections | HP:0002719 | 16%, (6/37) | 14%, (2/14) | 14%, (4/28) | 17%, (4/23) | 16%, (8/51) |
| | Recurrent otitis media | HP:0000403 | 8%, (3/38) | 7%, (1/14) | 7%, (2/29) | 9%, (2/23) | 8%, (4/52) |
| | Oral cleft | HP:0000202 | 5%, (2/38) | 0%, (0/15) | 7%, (2/29) | 0%, (0/24) | 4%, (2/53) |

**Table S2 | Literature reports of missense variants in other BAF subunits**

| PMID | DOI | Title |
|---|---|---|
| PMID:23637025 | 10.1002/ajmg.a.35933 | Clinical correlations of mutations affecting six components of the SWI/SNF complex: detailed description of 21 patients and a review of the literature |
| PMID:23661504 | 10.1002/aur.1301 | Quantifying repetitive speech in autism spectrum disorders and language impairment |
| PMID:23906836 | 10.1093/hmg/ddt366 | A comprehensive molecular study on Coffin-Siris and Nicolaides-Baraitser syndromes identifies a broad molecular and clinical spectrum converging on altered chromatin remodeling |
| PMID:26364901 | 10.1002/ajmg.a.37356 | Report of a patient with a constitutional missense mutation in SMARCB1, Coffin-Siris phenotype, and schwannomatosis |
| PMID:27264197 | 10.1002/ajmg.a.37722 | SMARCE1, a rare cause of Coffin-Siris Syndrome: Clinical description of three additional cases |
| PMID:30209809 | 10.1111/cge.13436 | New SMARCE1 variant in a patient with features overlapping with oculoauriculofrontonasal syndrome |
| PMID:30879640 | 10.1016/j.ajhg.2019.02.001 | A Syndromic Neurodevelopmental Disorder Caused by Mutations in SMARCD1, a Core SWI/SNF Subunit Needed for Context-Dependent Neuronal Gene Regulation in Flies |
| PMID:34706719 | 10.1186/s12920-021-01104-9 | Phenotypic and molecular spectra of patients with switch/sucrose nonfermenting complex-related intellectual disability disorders in Korea |
| PMID:35579625 | 10.1016/j.gim.2022.04.010 | Discovering a new part of the phenotypic spectrum of Coffin-Siris syndrome in a fetal cohort |
| PMID:35796094 | 10.1002/ajmg.a.62889 | Evidence for an association between Coffin-Siris syndrome and congenital diaphragmatic hernia |

**Table S3 | All truncating variants vs. truncating variants without carriers of c.3129del p.(Gly1044Aspfs*17)**

| Group | Phenotype | HPO | Truncating all | Truncating no c.3129 del | All cases | p-no3129delvT (nominal) | p-no3129delvT (FDR-corr.) | OR-no3129delvT (95% CI) |
|---|---|---|---|---|---|---|---|---|
| Intellectual and social development | Global developmental delay; Intellectual disability | HP:0001263; HP:0001249 | 72%, (21/29) | 87%, (20/23) | 85%, (47/55) | 0.3081120 | 0.308 (1.000) | 2.50 (0.51 - 16.69) |
| | mild GDD/ID | HP:0011342; HP:0001256 | 55%, (16/29) | 65%, (15/23) | 38%, (21/55) | 0.5729483 | 0.573 (1.000) | 1.51 (0.43 - 5.52) |
| | moderate/severe GDD/ID | HP:0011343; HP:0002342; HP:0011344; HP:0010864 | 17%, (5/29) | 22%, (5/23) | 47%, (26/55) | 0.7341153 | 0.734 (1.000) | 1.33 (0.26 - 6.74) |
| | Autistic behavior | HP:0000729 | 35%, (11/31) | 17%, (4/23) | 34%, (19/56) | 0.2199511 | 0.220 (1.000) | 0.39 (0.08 - 1.61) |
| | Behavioral abnormalities | HP:0000708 | 62%, (18/29) | 52%, (12/23) | 60%, (32/53) | 0.5755427 | 0.576 (1.000) | 0.67 (0.19 - 2.33) |
| Neurological system | Muscular hypotonia | HP:0001252 | 52%, (15/29) | 48%, (11/23) | 69%, (38/55) | 1.0000000 | 1.000 (1.000) | 0.86 (0.25 - 2.93) |
| | Brain imaging abnormality | HP:0410263 | 50%, (7/14) | 58%, (7/12) | 61%, (20/33) | 0.7126881 | 0.713 (1.000) | 1.38 (0.23 - 8.70) |
| | Visual impairment | HP:0000505 | 21%, (6/29) | 17%, (4/23) | 33%, (17/52) | 1.0000000 | 1.000 (1.000) | 0.81 (0.15 - 4.01) |
| | Seizures | HP:0001250 | 25%, (7/28) | 30%, (7/23) | 28%, (15/54) | 0.7572891 | 0.757 (1.000) | 1.31 (0.32 - 5.39) |
| | EEG abnormality | HP:0002353 | 20%, (4/20) | 27%, (4/15) | 23%, (7/30) | 0.7002730 | 0.700 (1.000) | 1.44 (0.22 - 9.59) |
| | Muscular hypertonia | HP:0001276 | 17%, (5/29) | 17%, (4/23) | 20%, (11/54) | 1.0000000 | 1.000 (1.000) | 1.01 (0.17 - 5.44) |
| | Hearing impairment | HP:0000365 | 10%, (3/29) | 4%, (1/23) | 11%, (6/53) | 0.6205818 | 0.621 (1.000) | 0.40 (0.01 - 5.40) |
| Craniofacial anomalies | Abnormality of the outer ear | HP:0000356 | 30%, (8/27) | 33%, (7/21) | 45%, (20/44) | 1.0000000 | 1.000 (1.000) | 1.18 (0.29 - 4.79) |
| | Thin upper lip vermilion | HP:0000219 | 48%, (14/29) | 48%, (11/23) | 43%, (23/53) | 1.0000000 | 1.000 (1.000) | 0.98 (0.29 - 3.36) |
| | Thick eyebrows | HP:0000574 | 38%, (11/29) | 39%, (9/23) | 38%, (20/53) | 1.0000000 | 1.000 (1.000) | 1.05 (0.29 - 3.72) |
| | Broad philtrum | HP:0000289 | 28%, (8/29) | 26%, (6/23) | 32%, (15/47) | 1.0000000 | 1.000 (1.000) | 0.93 (0.22 - 3.76) |
| | Prominent forehead | HP:0011220 | 26%, (7/27) | 19%, (4/21) | 32%, (14/44) | 0.7334165 | 0.733 (1.000) | 0.68 (0.12 - 3.23) |
| | Thick lower lip vermilion | HP:0000179 | 24%, (7/29) | 30%, (7/23) | 30%, (16/53) | 0.7550665 | 0.755 (1.000) | 1.37 (0.33 - 5.62) |
| | Short philtrum | HP:0000322 | 28%, (8/29) | 30%, (7/23) | 29%, (14/48) | 1.0000000 | 1.000 (1.000) | 1.15 (0.29 - 4.51) |
| | Long eyelashes | HP:0000527 | 21%, (6/29) | 26%, (6/23) | 29%, (15/52) | 0.7455236 | 0.746 (1.000) | 1.35 (0.30 - 6.04) |
| | Thick alae nasi | HP:0009928 | 24%, (7/29) | 30%, (7/23) | 27%, (14/51) | 0.7550665 | 0.755 (1.000) | 1.37 (0.33 - 5.62) |
| | Wide nose | HP:0000445 | 24%, (7/29) | 17%, (4/23) | 25%, (13/51) | 0.7353814 | 0.735 (1.000) | 0.67 (0.12 - 3.12) |
| | Down-slanting palpebral fissures | HP:0000494 | 21%, (6/29) | 22%, (5/23) | 21%, (10/48) | 1.0000000 | 1.000 (1.000) | 1.06 (0.22 - 4.97) |
| | Upturned nasal tip | HP:0000463 | 10%, (3/29) | 13%, (3/23) | 17%, (9/52) | 1.0000000 | 1.000 (1.000) | 1.29 (0.16 - 10.72) |
| | Wide mouth | HP:0000154 | 7%, (2/29) | 4%, (1/23) | 15%, (7/47) | 1.0000000 | 1.000 (1.000) | 0.62 (0.01 - 12.64) |
| | Hypertelorism | HP:0000316 | 10%, (3/29) | 13%, (3/23) | 14%, (7/49) | 1.0000000 | 1.000 (1.000) | 1.29 (0.16 - 10.72) |
| | Depressed nasal bridge | HP:0005280 | 10%, (3/29) | 13%, (3/23) | 12%, (6/51) | 1.0000000 | 1.000 (1.000) | 1.29 (0.16 - 10.72) |
| | Macrotia | HP:0000400 | 7%, (2/28) | 9%, (2/22) | 7%, (3/46) | 1.0000000 | 1.000 (1.000) | 1.29 (0.09 - 19.29) |

| Group | Phenotype | HPO | Truncating all | Truncating no c.3129 del | All cases | p-no3129delvT (nominal) | p-no3129delvT (FDR-corr.) | OR-no3129delvT (95% CI) |
|---|---|---|---|---|---|---|---|---|
| | Epicanthic fold | HP:0000286 | 3%, (1/29) | 4%, (1/23) | 6%, (3/48) | 1.0000000 | 1.000 (1.000) | 1.27 (0.02 -103.34) |
| | Up-slanting palpebral fissures | HP:0000582 | 0%, (0/29) | 0%, (0/23) | 4%, (2/48) | 1.0000000 | 1.000 (1.000) | 0.00 (0.00 - Inf) |
| | Downturned mouth | HP:0002714 | 3%, (1/29) | 4%, (1/23) | 2%, (1/48) | 1.0000000 | 1.000 (1.000) | 1.27 (0.02 -103.34) |
| Phenotypical abnormalities of body and face | Abnormal facial shape | HP:0001999 | 36%, (10/28) | 41%, (9/22) | 45%, (23/51) | 0.7741626 | 0.774 (1.000) | 1.24 (0.34 - 4.56) |
| | Abnormality of skeletal morphology | HP:0011842 | 26%, (7/27) | 29%, (6/21) | 33%, (17/51) | 1.0000000 | 1.000 (1.000) | 1.14 (0.26 - 4.93) |
| | Abnormality of the hand | HP:0001155 | 29%, (8/28) | 32%, (7/22) | 31%, (16/52) | 1.0000000 | 1.000 (1.000) | 1.16 (0.29 - 4.64) |
| | Decreased body weight | HP:0004325 | 11%, (3/27) | 14%, (3/21) | 30%, (15/50) | 1.0000000 | 1.000 (1.000) | 1.33 (0.16 - 11.10) |
| | Abnormality of the foot | HP:0001760 | 26%, (7/27) | 29%, (6/21) | 29%, (15/51) | 1.0000000 | 1.000 (1.000) | 1.14 (0.26 - 4.93) |
| | Abnormality of the eye | HP:0000478 | 8%, (2/26) | 5%, (1/20) | 29%, (14/48) | 1.0000000 | 1.000 (1.000) | 0.64 (0.01 - 13.13) |
| | Short stature | HP:0004322 | 7%, (2/27) | 10%, (2/21) | 24%, (12/50) | 1.0000000 | 1.000 (1.000) | 1.31 (0.09 - 19.58) |
| | Macrocephaly | HP:0000256 | 16%, (4/25) | 11%, (2/19) | 17%, (8/48) | 0.6842777 | 0.684 (1.000) | 0.62 (0.05 - 4.98) |
| | Microcephaly | HP:0000252 | 4%, (1/25) | 5%, (1/19) | 10%, (5/48) | 1.0000000 | 1.000 (1.000) | 1.32 (0.02 -108.87) |
| Skeletal anomalies | Scoliosis | HP:0002650 | 17%, (5/29) | 17%, (4/23) | 28%, (15/54) | 1.0000000 | 1.000 (1.000) | 1.01 (0.17 - 5.44) |
| | Joint laxity | HP:0001388 | 21%, (6/28) | 23%, (5/22) | 14%, (7/51) | 1.0000000 | 1.000 (1.000) | 1.08 (0.22 - 5.07) |
| | Delayed skeletal maturation | HP:0002750 | 6%, (1/16) | 9%, (1/11) | 9%, (3/33) | 1.0000000 | 1.000 (1.000) | 1.48 (0.02 -125.34) |
| | Clinodactyly | HP:0030084 | 7%, (2/29) | 9%, (2/23) | 8%, (4/52) | 1.0000000 | 1.000 (1.000) | 1.28 (0.09 - 19.02) |
| | Brachydactyly | HP:0001156 | 3%, (1/29) | 4%, (1/23) | 6%, (3/52) | 1.0000000 | 1.000 (1.000) | 1.27 (0.02 -103.34) |
| | Prominent interphalangeal joints | HP:0006237 | 4%, (1/27) | 5%, (1/21) | 4%, (2/51) | 1.0000000 | 1.000 (1.000) | 1.29 (0.02 -105.83) |
| | Pectus excavatum | HP:0000767 | 0%, (0/29) | 0%, (0/23) | 4%, (2/52) | 1.0000000 | 1.000 (1.000) | 0.00 (0.00 - Inf) |
| | Craniosynostosis | HP:0001363 | 4%, (1/28) | 0%, (0/22) | 2%, (1/51) | 1.0000000 | 1.000 (1.000) | 0.00 (0.00 - 49.60) |
| | Absent distal phalanges of 5th finger | HP:0005807 | 0%, (0/28) | 0%, (0/22) | 0%, (0/52) | 1.0000000 | 1.000 (1.000) | 0.00 (0.00 - Inf) |
| Ectodermal anomalies | Hypertrichosis | HP:0000998 | 14%, (4/29) | 17%, (4/23) | 17%, (9/54) | 1.0000000 | 1.000 (1.000) | 1.31 (0.21 - 8.01) |
| | Sparse/thin scalp hair | HP:0002209 | 7%, (2/29) | 9%, (2/23) | 16%, (9/55) | 1.0000000 | 1.000 (1.000) | 1.28 (0.09 - 19.02) |
| | Delayed eruption of primary teeth | HP:0000680 | 0%, (0/15) | 0%, (0/9) | 9%, (2/23) | 1.0000000 | 1.000 (1.000) | 0.00 (0.00 - Inf) |
| | Delayed eruption of permanent teeth | HP:0000696 | 0%, (0/12) | 0%, (0/6) | 5%, (1/19) | 1.0000000 | 1.000 (1.000) | 0.00 (0.00 - Inf) |
| | Aplasia/Hypoplasia of the nails | HP:0008386 | 0%, (0/29) | 0%, (0/23) | 4%, (2/52) | 1.0000000 | 1.000 (1.000) | 0.00 (0.00 - Inf) |
| | Microdontia | HP:0000691 | 0%, (0/25) | 0%, (0/19) | 0%, (0/34) | 1.0000000 | 1.000 (1.000) | 0.00 (0.00 - Inf) |
| Congenital anomalies | Abnormality of the gastrointestinal tract | HP:0011024 | 12%, (3/24) | 11%, (2/18) | 23%, (9/39) | 1.0000000 | 1.000 (1.000) | 0.88 (0.07 - 8.64) |
| | Abnormality of the genitourinary system | HP:0000119 | 13%, (3/23) | 6%, (1/17) | 22%, (8/37) | 0.6235037 | 0.624 (1.000) | 0.43 (0.01 - 5.89) |
| | Generalized abnormality of skin | HP:0011354 | 10%, (3/29) | 4%, (1/23) | 16%, (9/55) | 0.6205818 | 0.621 (1.000) | 0.40 (0.01 - 5.40) |

| Group | Phenotype | HPO | Truncating all | Truncating no c.3129 del | All cases | p-no3129delvT (nominal) | p-no3129delvT (FDR-corr.) | OR-no3129delvT (95% CI) |
|---|---|---|---|---|---|---|---|---|
| | Abnormal heart morphology | HP:0001627 | 11%, (3/28) | 9%, (2/22) | 12%, (6/49) | 1.0000000 | 1.000 (1.000) | 0.84 (0.06 - 8.06) |
| | Hernia | HP:0100790 | 4%, (1/27) | 5%, (1/21) | 12%, (6/51) | 1.0000000 | 1.000 (1.000) | 1.29 (0.02 -105.83) |
| | Abnormality of the kidney | HP:0000077 | 5%, (1/22) | 6%, (1/17) | 9%, (3/34) | 1.0000000 | 1.000 (1.000) | 1.30 (0.02 -107.77) |
| | Laryngotracheomalacia | HP:0008755 | 4%, (1/28) | 5%, (1/22) | 6%, (3/52) | 1.0000000 | 1.000 (1.000) | 1.28 (0.02 -104.52) |
| Miscellaneous | Feeding difficulties/failure to thrive | HP:0011968; HP:0001508 | 33%, (9/27) | 33%, (7/21) | 51%, (27/53) | 1.0000000 | 1.000 (1.000) | 1.00 (0.25 - 3.93) |
| | Sleep disturbance | HP:0002360 | 21%, (6/28) | 18%, (4/22) | 21%, (11/52) | 1.0000000 | 1.000 (1.000) | 0.82 (0.15 - 4.08) |
| | Recurrent infections | HP:0002719 | 14%, (4/28) | 14%, (3/22) | 16%, (8/51) | 1.0000000 | 1.000 (1.000) | 0.95 (0.12 - 6.37) |
| | Recurrent otitis media | HP:0000403 | 7%, (2/29) | 9%, (2/23) | 8%, (4/52) | 1.0000000 | 1.000 (1.000) | 1.28 (0.09 - 19.02) |
| | Oral cleft | HP:0000202 | 7%, (2/29) | 9%, (2/23) | 4%, (2/53) | 1.0000000 | 1.000 (1.000) | 1.28 (0.09 - 19.02) |

**Table S4 | Oligonucleotides used for In-Fusion mutagenesis of pCMV-BAF170-FLAG.**

| **Mutation** | **Orientation** | **Sequence** |
|---|---|---|
| c.230C>T (p.Pro77Leu) | Fw | 5'-TAAACTGCTGATCAAATGTTTCCTAGATTTCAAA-3' |
| | Rev | 5'-TTGATCAGCAGTTTAGTGAGCGGTGCA-3' |
| c.640A>G (p.Thr214Ala) | Fw | 5'-GTTACGACGCGTGGATCCCAGCGAGTG-3' |
| | Rev | 5'-TCCACGCGTCGTAACTGTCAGGATAGTAGCCC-3' |
| c.743T>C p.Phe248Ser | Fw | 5'-CGACACCTCCAATGAATGGATGAATGAGGAAGAC-3' |
| | Rev | 5'-TCATTGGAGGTGTCGGTGTCCAGGATC-3' |
| c.1327C>T (p.Arg443Trp) | Fw | 5'-CCATTGAGTGGAGGGCTCTCCCCGAGT-3' |
| | Rev | 5'-CCCTCCACTCAATGGCATGAACACTATTGT-3' |
| c.1919T>C p.(Leu640Pro) | Fw | 5'-GCATTTTCCTCGTCTTCCCATTGAAGACCCA-3' |
| | Rev | 5'-AGACGAGGAAAATGCAAGATGCACTCGTCCT-3' |
| c.2602G>C (p.Ala868Pro) | Fw | 5'-CCCTGGCCCCCGCCGCAGTGAAAGCTAAGC-3' |
| | Rev | 5'-CGGCGGGGGCCAGGGCGGCGGCAGC-3' |
| c.2678A>G p.(Glu893Gly) | Fw | 5'-GCTGGTGGGGACCCAGATGAAAAAGTTGGAGATC-3' |
| | Rev | 5'-TGGGTCCCCACCAGCAGGGCCACCAAAG-3' |

**Table S5 | Antibodies and dilutions used.**

| Antibody | Supplier & cat. no. | Method | Dilution |
|---|---|---|---|
| mouse monoclonal ARID1B | Abcam ab57461 | PLA | 1:70 |
| | | Western | 1:250 |
| mouse monoclonal BRG1 (SMARCA4) | Santa Cruz sc-17796 | PLA | 1:250 |
| | | Western | 1:1 000 |
| mouse monoclonal FLAG | Sigma F1804 | PLA/IF | 1:500 |
| | | Western | 1:10 000 |
| rabbit polyclonal FLAG | Sigma F7425 | PLA/IF | 1:500 |
| | | Western | 1:1 000 |
| rat monoclonal FLAG | Novus biologicals NBP1-06712SS | PLA/IF | 1:100 |
| rabbit polyclonal SMARCC1 | Cell Signaling #11956 | PLA | 1:250 |
| | | Western | 1:1 000 |
| rabbit polyclonal SMARCD1 | Santa Cruz sc-135843 | PLA | 1:500 |
| | | Western | 1:1000 |
| rabbit polyclonal SMARCE1 | betyhl laboratories A300-810A-T | PLA | 1:50 |
| | | Western | 1:1 000 |
| Cy3 conjugated goat anti rat | dianova | PLA/IF | 1:300 |

| | | | |
|---|---|---|---|
| | 112-165-003 | | |
| Cy3 conjugated goat anti mouse | dianova<br>115-165-146 | IF | 1:300 |
| HRP conjugated anti mouse | Invitrogen<br>G-21040 | Western | 1:10 000 |
| HRP conjugated anti rabbit | Cell Signaling<br>#7074 | Western | 1:3 000 |
| Tidy Blot reagent | BioRad<br>STAR209P | Western | 1:200 |

### SUPPLEMENTARY REFERENCES


1. Chen CA, Lattier J, Zhu W, et al. Retrospective analysis of a clinical exome sequencing cohort reveals the mutational spectrum and identifies candidate disease-associated loci for BAFopathies. *Genet Med Off J Am Coll Med Genet*. 2022;24(2):364-373. doi:10.1016/j.gim.2021.09.017
2. Li D, Downes H, Hou C, et al. Further supporting SMARCC2-related neurodevelopmental disorder through exome analysis and reanalysis in two patients. *Am J Med Genet A*. 2022;188(3):878-882. doi:10.1002/ajmg.a.62597
3. Sobreira N, Schiettecatte F, Valle D, Hamosh A. GeneMatcher: a matching tool for connecting investigators with an interest in the same gene. *Hum Mutat*. 2015;36(10):928-930. doi:10.1002/humu.22844
4. Machol K, Rousseau J, Ehresmann S, et al. Expanding the spectrum of BAF-related disorders: de novo variants in SMARCC2 cause a syndrome with intellectual disability and developmental delay. *Am J Hum Genet*. 2019;104(1):164-178. doi:10.1016/j.ajhg.2018.11.007
5. Yi S, Li M, Yang Q, et al. De novo SMARCC2 variant in a chinese woman with Coffin-Siris syndrome 8: a case report with mild intellectual disability and endocrinopathy. *J Mol Neurosci MN*. 2022;72(6):1293-1299. doi:10.1007/s12031-022-02010-0
6. Lo T, Kushima I, Aleksic B, et al. Sequencing of selected chromatin remodelling genes reveals increased burden of rare missense variants in ASD patients from the Japanese population. *Int Rev Psychiatry Abingdon Engl*. 2022;34(2):154-167. doi:10.1080/09540261.2022.2072193
7. Gofin Y, Zhao X, Gerard A, et al. Evidence for an association between Coffin-Siris syndrome and congenital diaphragmatic hernia. *Am J Med Genet A*. 2022;188(9):2718-2723. doi:10.1002/ajmg.a.62889
8. Sun H, Zhang S, Wang J, et al. Expanding the phenotype associated with SMARCC2 variants: a fetus with tetralogy of Fallot. *BMC Med Genomics*. 2022;15(1):40. doi:10.1186/s12920-022-01185-0
9. Xi Q, He W, Zhang XHF, Le HV, Massagué J. Genome-wide impact of the BRG1 SWI/SNF chromatin remodeler on the transforming growth factor β transcriptional program. *J Biol Chem*. 2008;283(2):1146-1155. doi:10.1074/jbc.M707479200
10. Wittmann MT, Katada S, Sock E, et al. scRNA sequencing uncovers a TCF4-dependent transcription factor network regulating commissure development in mouse. *Development*. 2021;148(14):dev196022. doi:10.1242/dev.196022
11. He S, Wu Z, Tian Y, et al. Structure of nucleosome-bound human BAF complex. *Science*. 2020;367(6480):875-881. doi:10.1126/science.aaz9761
12. Nixon KCJ, Rousseau J, Stone MH, et al. A syndromic neurodevelopmental disorder caused by mutations in SMARCD1, a core SWI/SNF subunit needed for context-dependent neuronal gene regulation in flies. *Am J Hum Genet*. 2019;104(4):596-610. doi:10.1016/j.ajhg.2019.02.001
13. Yano S, Fujimoto A, Morin-Leisk J, et al. New *SMARCE1* variant in a patient with features overlapping with oculoauriculofrontonasal syndrome. *Clin Genet*. 2018;94(5):487-488. doi:10.1111/cge.13436
